# Rates and predictors of data and code sharing in the medical and health sciences: A systematic review with meta-analysis of individual participant data

**DOI:** 10.1101/2023.03.22.23287607

**Authors:** Daniel G. Hamilton, Kyungwan Hong, Hannah Fraser, Anisa Rowhani-Farid, Fiona Fidler, Matthew J. Page

## Abstract

**Objectives:** Many meta-research studies have investigated rates and predictors of data and code sharing in medicine. However, most of these studies have been narrow in scope and modest in size. We aimed to synthesise the findings of this body of research to provide an accurate picture of how common data and code sharing is, how this frequency has changed over time, and what factors are associated with sharing.

**Design:** Systematic review with meta-analysis of individual participant data (IPD) from meta-research studies. Data sources: Ovid MEDLINE, Ovid Embase, MetaArXiv, medRxiv, and bioRxiv were searched from inception to July 1^st^, 2021.

**Eligibility criteria:** Studies that investigated data or code sharing across a sample of scientific articles presenting original medical and health research.

**Data extraction and synthesis:** Two authors independently screened records, assessed risk of bias, and extracted summary data from study reports. IPD were requested from authors when not publicly available. Key outcomes of interest were the prevalence of statements that declared data or code were publicly available, or ‘available on request’ (declared availability), and the success rates of retrieving these products (actual availability). The associations between data and code availability and several factors (e.g., journal policy, data type, study design, research subjects) were also examined. A two-stage approach to IPD meta-analysis was performed, with proportions and risk ratios pooled using the Hartung-Knapp-Sidik-Jonkman method for random-effects meta-analysis. Three-level random-effects meta-regressions were also performed to evaluate the influence of publication year on sharing rate.

**Results:** 105 meta-research studies examining 2,121,580 articles across 31 specialties were included in the review. Eligible studies examined a median of 195 primary articles (IQR: 113-475), with a median publication year of 2015 (IQR: 2012-2018). Only eight studies (8%) were classified as low risk of bias. Useable IPD were assembled for 100 studies (2,121,197 articles), of which 94 datasets passed independent reproducibility checks. Meta-analyses revealed declared and actual public data availability rates of 8% (95% CI: 5-11%, 95% PI: 0-30%, k=27, o=700,054) and 2% (95% CI: 1-3%, 95% PI: 0-11%, k=25, o=11,873) respectively since 2016. Meta-regression indicated that only declared data sharing rates have increased significantly over time. For public code sharing, both declared and actual availability rates were estimated to be less than 0.5% since 2016, and neither demonstrated any meaningful increases over time. Only 33% of authors (95% CI: 5-69%, k=3, o=429) were estimated to comply with mandatory data sharing policies of journals.

**Conclusion:** Code sharing remains persistently low across medicine and health research. In contrast, declarations of data sharing are also low, but they are increasing. However, they do not always correspond to the actual sharing of data. Mandatory data sharing policies of journals may also not be as effective as expected, and may vary in effectiveness according to data type - a finding that may be informative for policymakers when designing policies and allocating resources to audit compliance.

---

Data collection, analysis, and curation, each play integral roles in the research lifecycle across most scholarly fields, including medicine and health. It is also well recognised that archived research products like raw data and analytic code are valuable commodities to the broader medical research community. Among other things, greater access to raw data, analytic code and other materials that underly research findings provides researchers with opportunities to strengthen their methods, validate discovered findings, answer questions not originally considered by the data creators, accelerate research through the synthesis of existing datasets, and educate new generations of medical researchers [1]. While there are many valid challenges with sharing research materials (particularly navigating privacy considerations and time and resource burdens), in recognition of the benefits, funders and publishers of medical research have been carefully and continuously increasing the pressure on medical researchers over the last two decades to maximise the availability of such products for other researchers [2-6]. Recent examples include the United States government advising its federal funding agencies to update their public access policies before the end of 2025 to require that all federally funded research publications and supporting data are freely and immediately available [7].

While policy changes have fuelled optimism that data and code sharing rates in medicine will increase, important questions remain around what the culture of sharing is like currently, how it has evolved over time, how successful stakeholder policies are at instigating sharing, and when researchers are observed to share, how often useful data are made available. Many meta-research studies in medicine have aimed to address these questions, however, most have been small in size and narrow in scope, focussing on specific research participants (e.g., human participants [8], animals [9]), data types (e.g., gene expression data [10], modelling data [11]), study designs (e.g., clinical trials [12], systematic reviews [13]), and outcomes (e.g., data and code sharing declarations [14], data ‘FAIRness’ [15]). Therefore, the objectives of this review are to synthesise the findings of this research to establish an accurate picture of how common data and code sharing is in medicine, assess compliance with stakeholder policies on data and code availability, as well as explore what factors are associated with sharing. We anticipate that the findings of this review will highlight several areas for future policymaking and meta-research activities.

## METHODS

### Protocol and registration

We registered our systematic review on May 28th, 2021 on the Open Science Framework (OSF), prior to commencing the literature search [16], and subsequently prepared a detailed review protocol [17]. We report seven deviations from the protocol in Supplementary Table 1. As the research subjects of interest were scientific publications, ethics approval was not required for this research. The findings of this review are reported in accordance with the Preferred Reporting Items for Systematic reviews and Meta-Analyses (PRISMA) 2020 statement [18] and its IPD extension [19]. We summarise key aspects of the methods below; for further details, please refer to the review protocol [17].

### Eligibility criteria

Any study in which researchers investigated the prevalence of, or factors associated with, data or code sharing (termed “meta-research studies”) across a sample of published scientific articles presenting original medical or health-related research findings (termed “primary articles”) was eligible for inclusion in the review. No restrictions were placed on the publication location (e.g., preprint server, peer-reviewed journal) or the format (e.g., conference abstract, research letter) of either group. Nor were restrictions placed on the strategy used to identify and select primary articles, the type of data assessed (e.g., trial data, review data) or the level of sharing assessed (e.g., partial versus complete sharing). Furthermore, we included studies that used either manual or automated methods to assess data and code sharing provided it involved some examination of the body text of sampled primary articles. Exclusion criteria for this review included meta-research studies that investigated data or code sharing: as a routine part of a systematic review and IPD meta-analysis; among scientific articles outside of medicine and health; or via avenues other than journal articles (e.g., clinical trial registries).

### Information sources and search strategy

On July 1st, 2021, we searched Ovid MEDLINE, Ovid Embase, and the medRxiv, bioRxiv, and MetaArXiv preprint servers to identify potentially relevant studies indexed from database inception up to the search date. The full search strategies, bibliographic citation files, as well as snapshots of the medRxiv and bioRxiv databases are available on the project’s OSF page [20]. Details on the development of the search strategy are outlined in the review protocol [17]. In addition to the database searches, other preprint servers (PeerJ, Research Square) and relevant online resources (Open Science Framework, aspredicted.org and connectedpapers.com) were searched to locate additional published, unpublished and registered studies of relevance to the review. Backward and forward citation searches of meta-research studies meeting the inclusion criteria were also performed using citationchaser on August 30th, 2022 [21]. Finally, potentially relevant studies recommended by colleagues, discovered through collaborations, and seen at meta-research conferences were also screened for eligibility. No language restrictions were imposed on any of the searches.

### Study selection

Results from all main database and preprint server searches were imported into Covidence (Covidence systematic review software, Veritas Health Innovation, Melbourne, Australia) and deduplicated. For the preprint searches, if a version of an eligible meta-research study was discovered in a peer-reviewed journal, it was included in place of the original preprint. All titles, abstracts, and full-text articles were then screened for eligibility in Covidence by DGH and another author (HF, ARF, or KH) independently, with disagreements resolved via discussion between authors, or by a third author if necessary (MJP). All literature identified by the additional preprint and online searches were screened against the eligibility criteria by one author (DGH). When multiple reports on the same dataset were identified, we used data from the most up-to-date report. A spreadsheet containing all screening decisions is available on the project’s OSF page [20].

### Data collection

Once a meta-research study was found to be eligible, one member of the team (DGH) determined whether sufficiently unprocessed article-level IPD and article identifiers (e.g., digital object identifiers (DOIs), PubMed identifiers (PMIDs), article titles) for the included primary articles were publicly available. For meta-research studies where complete IPD were not available (i.e., no data or partial data had been shared), the corresponding author was contacted and asked if they would provide the complete or remaining IPD. If meta-research authors responded that they were either unable or unwilling to share, we then asked whether they would calculate the summary statistics necessary for the review. For meta-research authors who were unable or refused to provide summary data for the review, did not respond, or did not provide the promised IPD by the census date of December 31st, 2022, summary data reported in the meta-research papers were independently extracted by two authors (DGH; MJP), with discrepancies resolved through discussion. A list of all the data that were extracted from each meta-research study for the review can be found on the project’s OSF page [20].

### Assessments of risk of bias

The risk of bias of included meta-research studies was assessed using a tool designed based on methods used in previous Cochrane Methodology reviews [22, 23]. The tool included four domains: i) sampling bias, ii) selective reporting bias, iii) article selection bias, and iv) the risk of errors in the accuracy of reported estimates (Supplementary Table 2). Each meta-research article was independently assessed by DGH and one other author (KH or ARF), with discrepancies resolved via discussion, or a third author (MJP) if necessary. Where domains were rated as unclear, clarification was sought from meta-research authors. Given the purpose of the tool was to differentiate between studies at a high risk of bias from those with a low risk, a study was only classified as low risk of bias if all criteria were assessed as low risk. We did not assess the likelihood of publication bias affecting the findings of the review (e.g., using a funnel plot), nor did we assess certainty in the body of evidence, as available methods are not well suited for methodology reviews such as ours.

### IPD integrity checks and harmonisation

When complete IPD were obtained for a meta-research study, one author (DGH) performed the following integrity checks prior to harmonising the data: i) an evaluation of the completeness of the dataset (e.g., whether any variables or values were missing), ii) a check of the validity of the dataset (e.g., presence of out-of-range values, incorrectly coded values) and iii) a check that the overall sample size and data and/or code sharing rates as stated in the report could be exactly reproduced (note that the checks for an included study led by the first author of this review (Hamilton et al 2022 [15]) were performed by another author (HF)). In instances where any of these checks failed, clarification was sought from the meta-research authors. We also checked for, and removed duplicate rows in datasets (i.e., checked if the same primary articles were sampled more than once). Additionally, for meta-research studies that sampled primary articles across multiple scientific disciplines, Digital Science’s Dimensions platform (https://app.dimensions.ai) was used to identify which were medical and health-related using their automated 2020 Australia and New Zealand Standard Research Classification (ANZSRC) Fields of Research (FOR) Codes classification service [24]. When primary articles were not indexed in Dimensions, the first author (DGH), who has close to a decade of experience working as an allied health professional, clinical trial coordinator and medical researcher, classified articles as being medical or health-related or not. Furthermore, for meta-research studies with sample sizes less than 500, primary articles not assigned medical FOR codes by the Dimensions platform were manually reviewed and recoded if deemed false negatives.

Once the IPD checks were complete, one author (DGH) then manually extracted and reclassified required data in line with the study’s codebook. When all available IPD had been assembled and harmonised, datasets were then merged and the extent of overlapping primary articles between meta-research studies was assessed for each outcome of interest by checking for duplicate DOIs and PMIDs in R (R Foundation for Statistical Computing, Vienna, Austria, v4.2.1) using the duplicated function. We decided to keep data originating from primary articles that were flagged as having been sampled by more than one meta-research study only for the study with the highest score for the fourth risk of bias domain (i.e., lowest risk of errors in the accuracy of reported estimates), or in the event of a tie, the overall lowest risk of bias judgement, or the most recent publication date. More details on the scoring system developed to resolve overlap can be found on the project’s OSF page. For eligible meta-research studies where summary data were only available from study reports, but primary study identifiers were known, information from overlapping primary articles was removed from the meta-research studies that shared complete IPD. For meta-research studies where both primary study identifiers and article-level data were unavailable, we assessed the likelihood of overlap with other meta-research studies by comparing: i) outcome data collected, ii) primary article date range and iii) sampled journals.

### Outcomes of interest

The following four pre-specified outcome measures for both research data and code availability were of primary interest to the review:

i. the prevalence of primary articles where authors declared that their data or code are publicly available (‘declared public availability’);
ii. the prevalence of primary articles in which meta-researchers verified that data or code were indeed publicly available (‘actual public availability’);
iii. the prevalence of primary articles where authors declared their data or code are privately available (i.e., “available on request” statements) (‘declared private availability’), and;
iv. the prevalence of primary articles in which meta-researchers confirmed that study data or code were released in response to a private request (‘actual private availability’).

‘Actual public availability’ represented the results of the most intensive investigation of an availability statement by meta-researchers (e.g., checks that reported URLs were functional, that data could befreely downloaded and opened, that datasets were complete, that reported results could be independently reproduced). We also required data to be immediately available for it to be classified as actually publicly available (i.e., did not accept ‘intention to share’ and ‘under embargo’ statements), and took the strictest definition of actual availability when alternatives were available (i.e., if a study assessed both partial and complete sharing, we took the results of the ‘full’ data availability). Further information on how we defined ‘actual availability’ as well as all our other outcome measures can be found in the review protocol and the study codebook on the project’s OSF page [20].

In addition to the primary outcome measures, we also included eight secondary outcome measures:

i. the prevalence of formalised sections within primary articles dedicated to addressing data and/or code availability;
ii. the association between the presence of a data availability statement and public sharing of data in primary articles;
iii. the association between the presence of a code availability statement and public sharing of research code in primary articles;
iv. the association between a journal’s policy on data sharing (any ‘mandatory posting’ policy versus other policy) and public sharing of research data in primary articles;
v. the association between a journal’s policy on data sharing (‘make available on request’ policy versus other non-mandatory policy) and private sharing of research data in primary articles;
vi. the association between study design (clinical trial versus non-trial) and public sharing of data in primary articles;
vii. the association between the subjects of the research (human participants versus non-human participants) and public sharing of data in primary articles, and;
viii. the association between public sharing of research data and the sharing of code in primary articles.

## Statistical analysis

A ‘two-stage’ approach to IPD meta-analysis was used, whereby summary statistics were computed from available IPD, abstracted from included study reports, or obtained directly from meta-research authors, then pooled using conventional meta-analysis techniques. We calculated proportions and 95% confidence intervals (CI) for all prevalence outcomes. Where possible, we calculated risk ratios with 95% confidence intervals for all association outcomes. For primary outcome measures, we considered the methodological characteristics of the included studies to determine which were appropriate for aggregation and decided that we would pool studies that met the following criteria: i) did not use non-random sampling methods, ii) did not restrict primary article evaluations to specific journals, preprint servers, funders, institutions, or data types, and iii) reported outcome data on primary articles published after 2016. These criteria were specifically chosen to minimise biasing of estimates (i.e., reduce upward or downward biasing of pooled estimates due to the overrepresentation of studies of journals with mandatory sharing policies, certain study designs, etc), and to provide a modern picture of data and code sharing (i.e., an estimate of sharing since the introduction of the FAIR principles [25]). The same criteria were applied to secondary outcome measures and subgroup analyses unless specified otherwise.

We pooled prevalence estimates by first stabilising the variances of the raw proportions using arcsine square root transformations, then applied random-effects models using the Hartung-Knapp-Sidik-Jonkman method which has shown to be preferable to the DerSimonian and Laird method when including a small number of studies, and when including studies with differing sample sizes [26]. The same approach was also used for meta-analyses of risk ratios; however, no transformations were used, and the ‘treatment arm’ continuity correction proposed by Sweeting et al 2004 [27] was applied to studies reporting zero events in a single group (double zero-cell events were excluded from the main analysis). Statistical heterogeneity was assessed via visual inspection of forest plots, the size of the I^2^ statistics and their 95% confidence intervals, and via 95% prediction intervals (PI) where more than four studies were included. Data deduplication, preparation, analysis and visualisation was performed in R (R Foundation for Statistical Computing, Vienna, Austria, v4.2.1) using the meta (v5.5) [28], metafor (v3.8) [29] and altmeta (v4.1) [30] packages. Risk of bias plots were created using robvis [31]. The Python (v3.10.7) client Dimcli (v0.9.9.1) was used to access Dimensions Analytic’s API and retrieve required primary article meta-data (e.g., DOIs, PMIDs, ANZSRC FOR codes). All R and Python scripts are publicly available on the project’s OSF page [20].

### Subgroup and sensitivity analyses

We planned to conduct the following subgroup analyses to investigate whether prevalence estimates of public data sharing differed depending on i) the data type, or whether primary articles: ii) were subject to any mandatory sharing policies by the funders of the research or not, or iii) posted a preprint prior to publication or not. Furthermore, we also investigated the influence of publication year on data and code sharing rates by fitting three-level mixed-effects meta-regression models on arcsine-transformed proportions. A multi-level model was used to account for dependencies between effect estimates due to some studies contributing multiple yearly estimates. Due to substantially differing levels of variation between the pre- and post-2014 periods, to preserve assumptions of homoscedasticity we only modelled changes in sharing rates from 2014 onwards.

We also performed sensitivity analyses to assess changes in pooled estimates when excluding meta-research studies that i) were rated as high or unclear risk of bias, ii) did not provide IPD for the review, iii) were at high risk of overlap with other meta-research studies, iv) did not assess compliance with the FAIR principles, v) did not manually assess primary articles and vi) did not examine COVID-19-related research. Finally, we also examined differences in pooled proportions and risk ratios when using generalised linear mixed models (GLMMs) to aggregate findings, which have been specifically recommended in situations when the probability of the event of interest is rare [32,33]. Such methods also circumvent the need to add arbitrary continuity corrections to zero events, which can produce biased results when most cases are zero events, and group sample sizes are highly imbalanced [27]. For meta-analyses of risk ratios, we report the results of analyses both excluding and including studies with no events in both groups.

## RESULTS

### Study selection and IPD retrieval

The search of Ovid MEDLINE, Ovid Embase and the medRxiv, bioRxiv and MetaArXiv preprint servers, once deduplicated, identified 4,952 potentially eligible articles for the review, of which 4,736 were excluded following the screening of titles and abstracts. Of the remaining 216 articles, full-text articles were retrieved for all papers, and 70 were adjudicated as eligible for the review. Furthermore, the additional searches revealed another 44 eligible reports for inclusion, resulting in a total of 114 eligible meta-research studies examining a combined total of 2,254,031 primary articles for the review [8-15,34-142]. Following confirmation of eligibility, we searched for publicly available IPD for the 114 meta-research studies. Of these studies, 70 had already made complete IPD publicly available (61%), 20 studies had posted partial IPD (18%), and 24 had not publicly shared any IPD (21%), with three of the latter articles declaring upfront that IPD could not be shared. Of the 70 complete datasets that were originally posted publicly, 60 (86%) were deposited into data repositories, 36 (51%) had a DOI, 26 (37%) provided a data dictionary, and 14 (20%) applied a license to the data. Most data were archived in Microsoft Excel (N=33, 47%) or CSV (N=25, 36%) formats, with a minority of meta-researchers storing their data in PDFs (N=5, 7%) and Microsoft Word documents (N=3, 4%).

Of the 40 meta-research studies that had not posted complete IPD, did not state in the study report that data could not be released, and had publicly available contact information, we contacted all authors and asked them to share article-level IPD for the review. We received 32 responses to our 40 requests (80%), of which 20 meta-researchers (50%) shared the required IPD by the census date. The median time taken to receive IPD was 7 days (range: 0-216 days). For the 20 articles where complete IPD was not assembled, 10 studies had useable IPD and/or summary data. The nine studies that were eligible for the review but could not be included in the quantitative analysis are outlined in Supplementary Table 3. They are also included in relevant forest plots, without providing usable data for the meta-analysis. Ultimately, 108 reports of 105 meta-research studies collecting information from a total of 2,121,580 primary articles were included in the quantitative analysis [8-15,34-133], with complete IPD available for 90 studies, a combination of partial IPD and summary data for 10 studies, and only summary data available for 5 studies. Refer to Figure 1 for the full PRISMA flow diagram.

**Figure 1.**
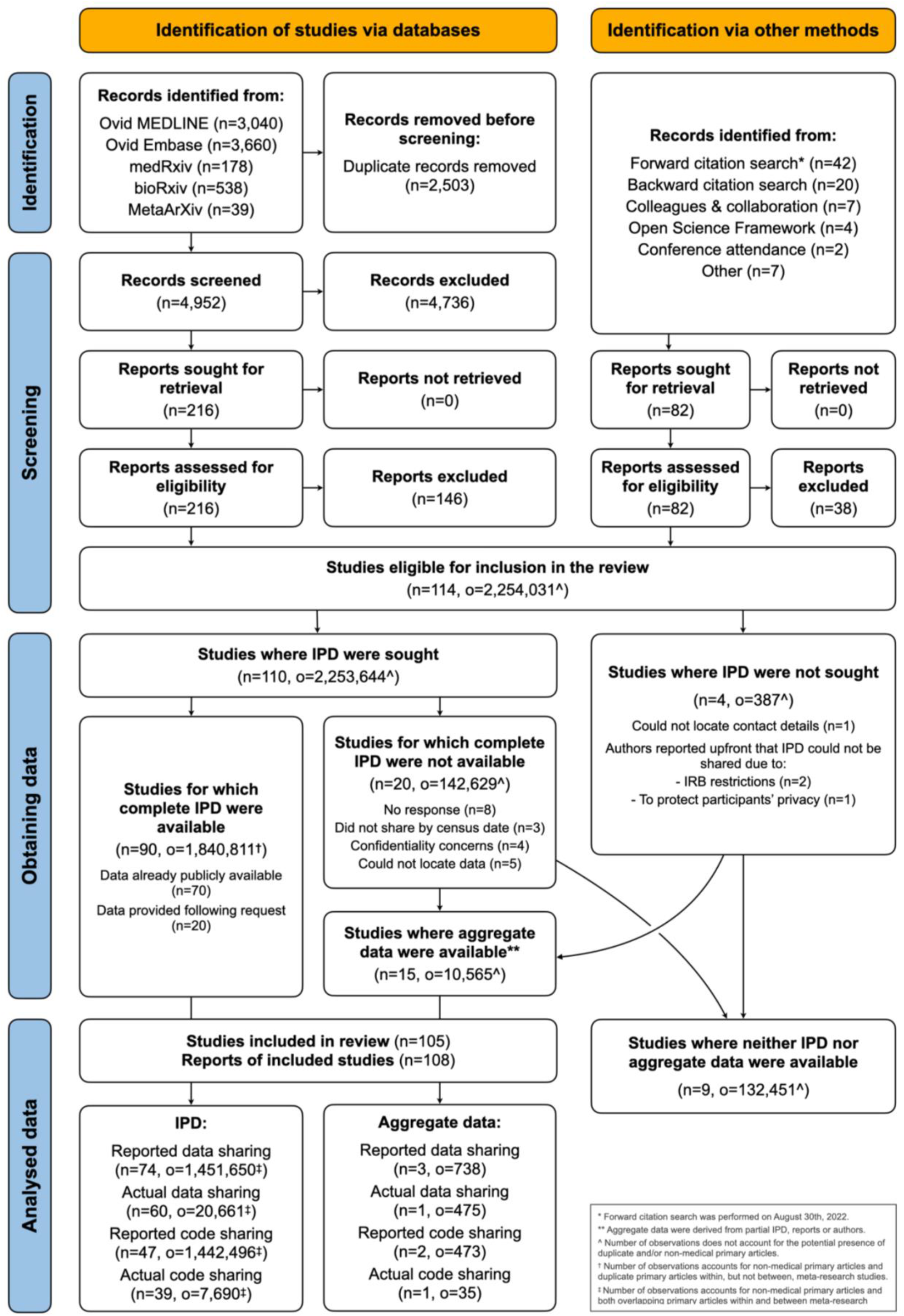
PRISMA 2020 and PRISMA-IPD flow diagram.

### Study characteristics

Summary information on the 105 meta-research studies that are included in the quantitative analysis of this review is outlined in Table 1. Eligible meta-research studies examined a median of 195 primary articles (IQR: 113-475; sample size range: 10-1,475,401), with a median publication year of 2015 (IQR: 2012-2018, publication date range: 1781-2022). Meta-research studies assessed data and code sharing across 31 specialties. Most commonly, studies were interdisciplinary, examining several medical fields simultaneously (N=17, 16%), followed by biomedicine and infectious disease (each N=10, 10%), general medicine (N=9, 9%), addiction medicine, clinical psychology, and oncology (each N=5, 5%). Eleven studies (10%) examined COVID-19-related articles. Additionally, most meta-research studies did not set any restrictions concerning data types (N=63) or journals of interest (N=56). However, when data restrictions were imposed, they were most often limited to trial data (N=16), sequence data (N=6), gene expression data and review data (each N=5). When journal restrictions were incorporated, the scope was most often limited to papers published in ‘high impact’ journals (variably defined by authors) (N=18), one or two journals of interest (N=10 and 5 respectively), or multiple journals subjectively deemed relevant to a field (N=7). Of the 105 meta-research studies, 31 and 4 also evaluated compliance with journal data and code sharing policies, respectively. However, none of the meta-research studies examined compliance with policies instituted by medical research funders or institutions.

**Table 1.**
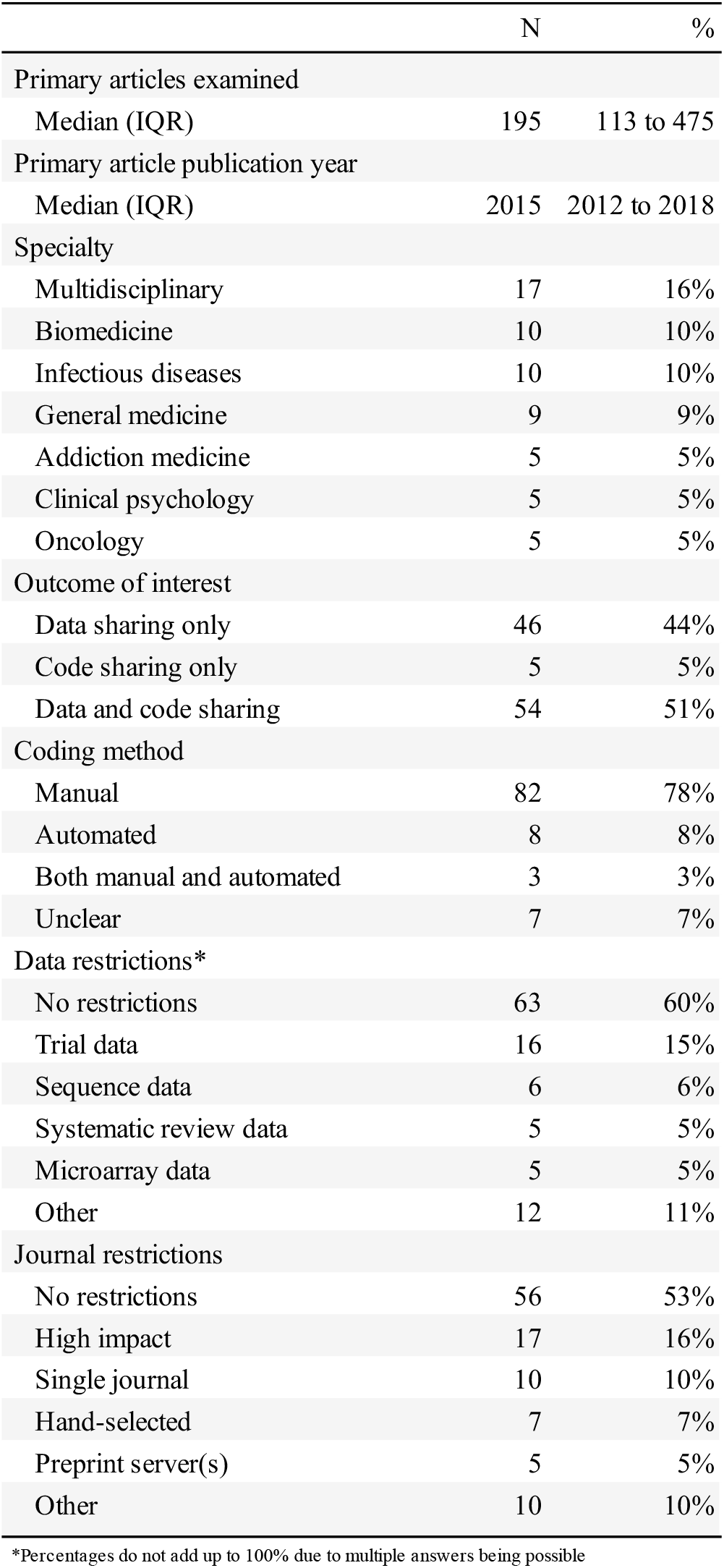
Characteristics of the included studies (N=105). The underlying data can found be at https://osf.io/ca89e.

In total, 95 and 58 meta-research studies, respectively, examined the prevalence of public data and code sharing in primary articles, with five studies examining how compliant publicly shared data was with the FAIR principles. In contrast, 10, 4 and 2 studies, respectively, assessed whether study data, code, or both data and code could be retrieved in response to a private request (i.e., actual private availability). Of these 16 studies, the stated reasons underpinning requests were: to perform a re-analysis (N=6), for a meta-research study (N=5), to populate a registry (N=1), to validate their findings (N=1) and for interest and coursework (N=1), with the remaining two not reporting what reason they gave. Of the 14 meta-research articles that shared the request templates they used, 12 meta-researchers provided primary article authors with an honest account of why they wished to source data and/or code, whereas two used deception.

### Risk of bias assessment

The overall and individual results of the risk of bias assessments are reported in Supplementary Figures 1 & 2. Most eligible meta-research studies were judged favourably on the first risk of bias domain (sampling bias), having randomly sampled primary articles from populations of interest, or assessed all eligible articles identified by their literature searches (N=95, 90%). In contrast, a minority of meta-research studies were judged to be at low risk of selective reporting bias (N=45, 42%) and article selection bias (N=24, 23%) (i.e., shared study protocols and information on which primary articles were excluded and why). Similarly, only half of meta-research studies (N=54, 51%) were judged to have used a primary article coding strategy considered to be at low risk of errors. Ultimately, only eight studies (8%) were classified as low risk of bias for all four domains.

### IPD integrity checks

In total, 100 meta-researchers’ datasets (90 complete and 10 partial) were obtained for the review. For the 90 complete datasets, sample sizes, as well as data and/or code sharing rates reported in study reports, were reproduced in all but five cases (94%), with the reasons for irreproducibility being due to simple typographical errors in the report (N=2), unclear data filtering steps (N=2) and an error in the meta-researchers’ code (N=1). For the ten partial datasets, we were able to independently verify sample sizes and sharing estimates for all but one case due to the receipt of an incorrect version of the data.

Of the 105 included meta-research studies examining 2,121,580 primary articles, we were able to retrieve identifying details (i.e., DOIs, PMIDs) for 2,121,197 primary articles (99.98%) from 100 studies (95%). After the removal of non-medical articles and duplicate articles observed within each of the 100 datasets, we were left with 1,849,828 primary articles with which to explore the extent of overlap between eligible studies. Of these 1,849,828 primary articles, we observed that 704,310 (38%) were flagged as having been sampled by more than one included meta-research study (some articles being repeatedly sampled by up to five studies). Notably, articles examined by the three largest studies by Serghiou et al [14], Colavizza et al [43] and Federer et al [50] were implicated in 681,595 of the 704,310 flagged cases (96.77%). Further, for some studies, all sampled primary articles had been completely assessed by other included studies (e.g., Sumner et al [122], Strcic et al [121]), whereas others demonstrated zero overlap (e.g. Rufiange et al [9]) (see Supplementary Figure 3 for further details).

For the five meta-research studies where identifying details for the primary articles were unavailable, only a single study was deemed to be at high risk of overlap [73]. Furthermore, for the nine meta-research studies excluded from the quantitative analysis, 127,985 of the 132,451 observations (97%) would have come from two meta-research studies of articles published in PLOS One, which would have had a high risk of overlap with the included studies by Serghiou et al [14], Colavizza et al [43] and Federer et al [50]. Given the likelihood of high overlap, our inability to include these nine meta-research studies in the quantitative analyses is unlikely to have influenced our results.

### Public data and code sharing rates

Combination of eligible studies in a random-effects meta-analysis suggests that 8% of medical articles published since 2016 declare data to be publicly available (95% CI: 5-11%, 95% PI: 0-30%, k = 27 studies, o = 700,054 primary articles, I^2^ = 96%; Figure 2) and 2% actually share data publicly (95% CI: 1-3%, 95% PI: 0-11%, k = 25, o = 11,873, I^2^ = 90%; Figure 3). Despite the included meta-research studies following similar methodologies, we do note high statistical heterogeneity for both analyses, with influence analyses showing that the greatest contributors to between-study heterogeneity for declared data sharing were the high precision findings of Uribe et al [125] and Serghiou et al [14], who used automated coding strategies. For actual data sharing, the high estimate by Hamilton et al [15], who assessed partial sharing of data rather than complete, was also a large contributor to between-study heterogeneity.

**Figure 2.**
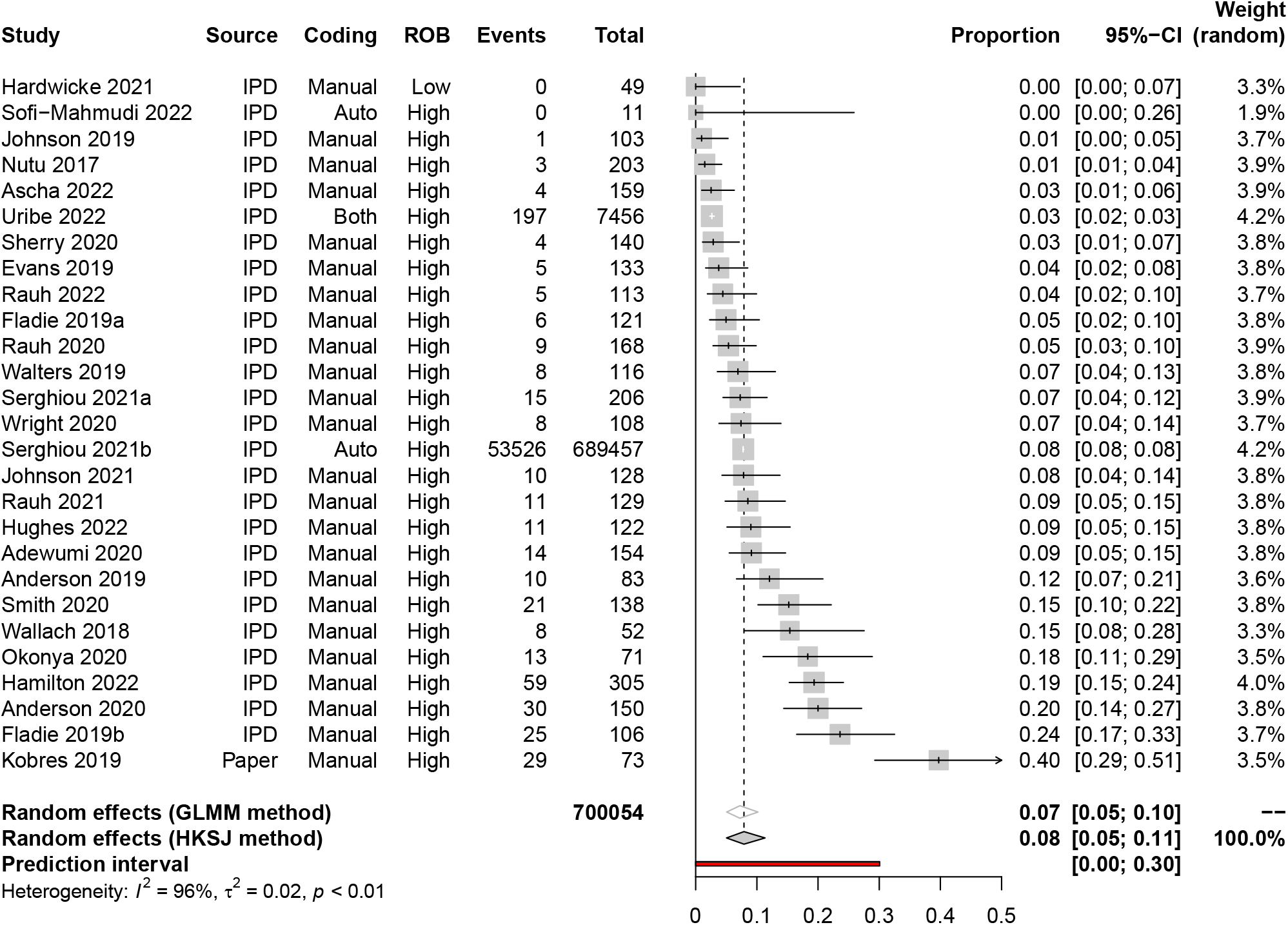
Declared public data sharing rates since 2016.

**Figure 3.**
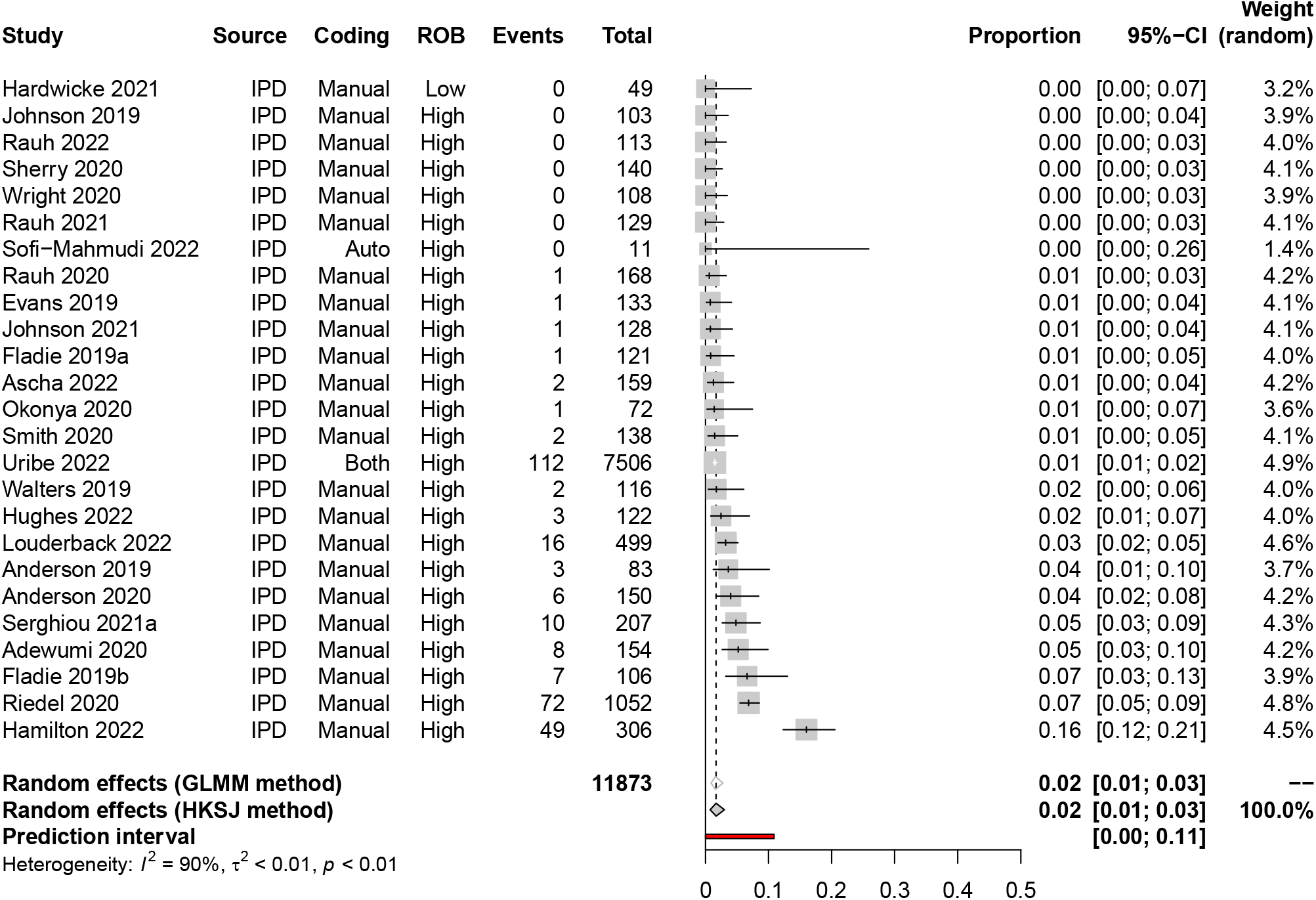
Actual public data sharing rates since 2016.

For public code sharing, declared and actual code sharing rates since 2016 are estimated to be 0.3% (95% CI: 0-1%, 95% PI: 0-8%, k = 26, o = 707,943, I^2^ = 89%; Figure 4) and 0.1% (95% CI: 0-0.3%, 95% PI: 0-1%, k = 21, o = 3,843, I^2^ = 52%; Figure 5), respectively. Like declared data sharing rates, despite similar methodologies, declared code sharing estimates were also associated with high statistical heterogeneity. Again, influence analyses revealed high precision estimates from Uribe et al [125] and Serghiou et al [14], in addition to the high estimate by Kobres et al [78], who evaluated the sharing of model code from Zika virus forecasting and prediction research, were the biggest contributors to between-study heterogeneity.

**Figure 4.**
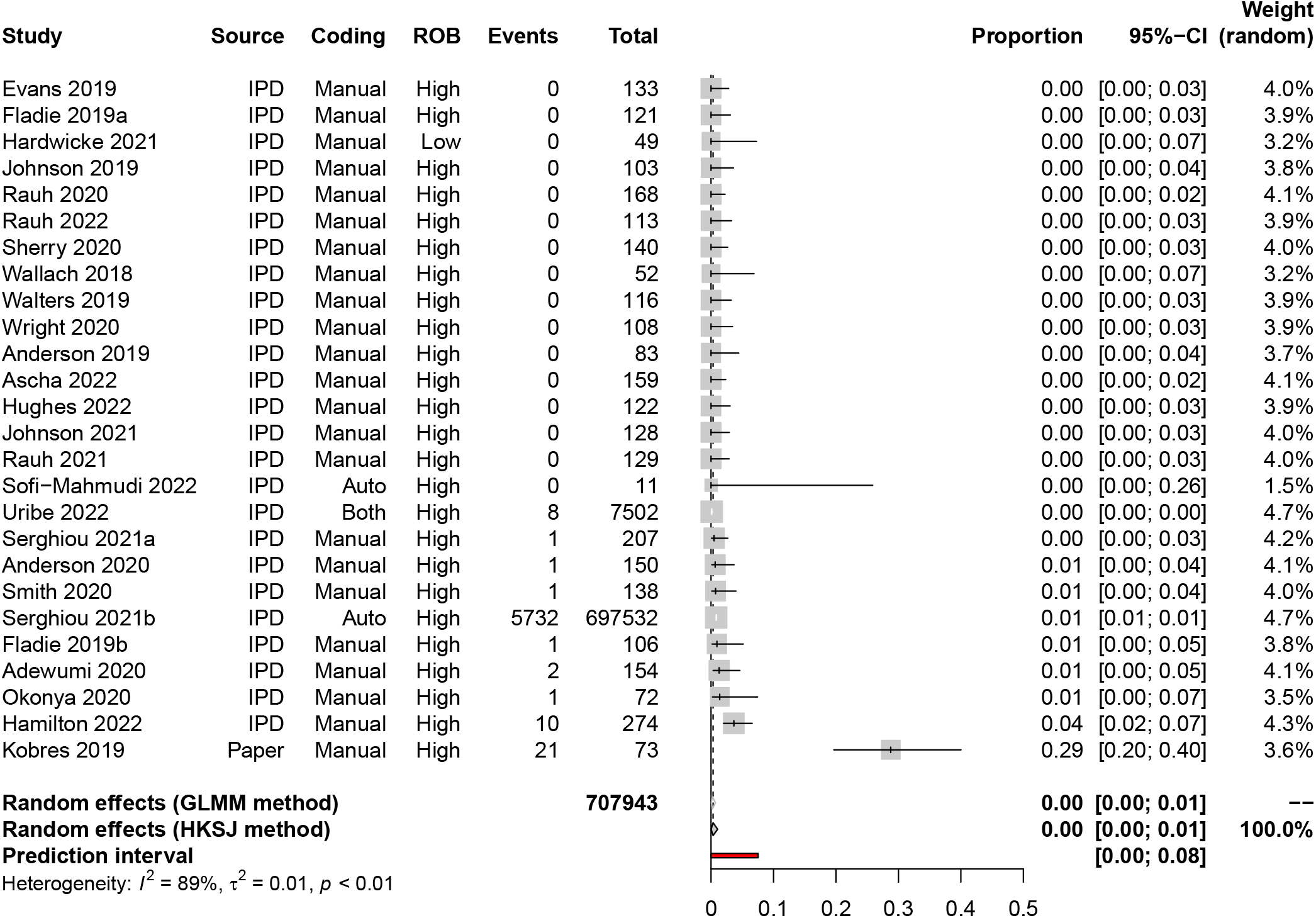
Declared public code sharing rates since 2016.

**Figure 5.**
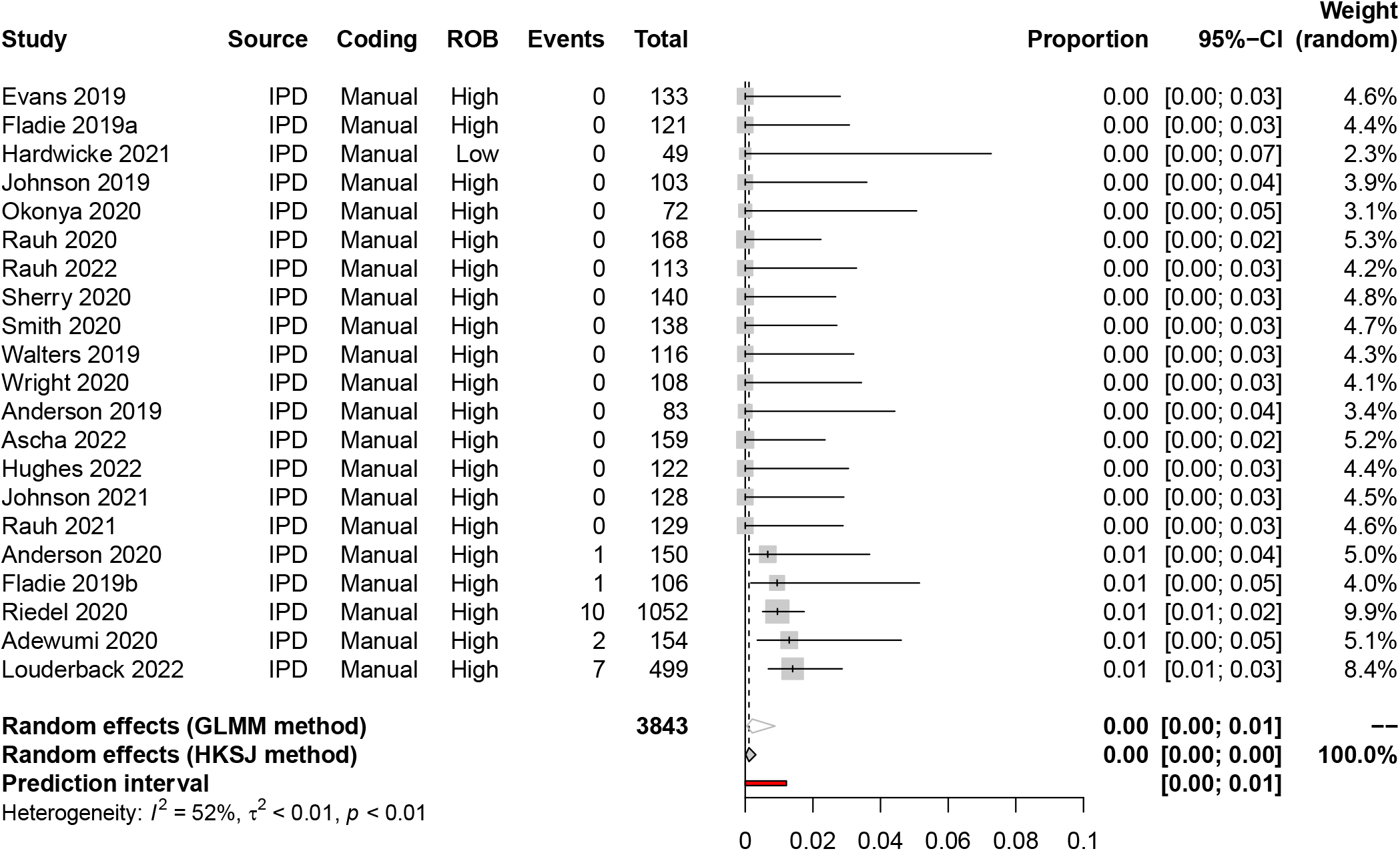
Actual public code sharing rates since 2016.

### Private data and code sharing rates

In contrast to declarations of public availability, ‘available upon request’ declarations were not commonly observed in primary articles published since 2016 for data (2%, 95% CI: 1-4%, 95% PI: 0-10%, k = 23, o = 3,058, I^2^ = 80%) or code (0%, 95% CI: 0-0.1%, 95% PI: 0-0.5%, k = 22, o = 2,825, I^2^ = 0%) (refer to Supplementary Figures 4 & 5 for forest plots). For actual private data and code availability rates, we could not combine the findings of eligible meta-research studies due to methodological differences, particularly in journal restrictions (i.e., policy differences), as well as the type of data being requested, both of which are explored via subgroup analyses below.

Overall, we observed that success rates in privately obtaining data and code from authors of published medical research ranged between 0-37% (k = 12, I^2^ = 88%) and 0-23% (k = 5, I^2^ = 94%) respectively (Figure 6). However, we note that when authors who declared data and code to be ‘available on request’ were asked for these products by meta-researchers, the upper limits of success increased to 100% (k = 7, I^2^ = 83%) and 43% (k = 4, I^2^ = 86%) respectively. In comparison, when requests for data and code were made to authors who did not include a statement concerning availability, success rates dropped to between 0-30% (k=7, I^2^ = 65%) and 0-12% (k=3, I^2^ = 89%) respectively. Lastly, and unsurprisingly, we also note that attempts to obtain data from authors explicitly declaring it to be unavailable were associated with a 0% sharing rate (k = 2, I^2^ = 0%). See Supplementary Figure 6 for the full results. Interestingly, we also noted during the IPD deduplication process that two of four primary article authors who were asked to share data by two independent meta-research teams on two separate occasions responded differently, providing some anecdotal evidence that requestor and requestee characteristics likely also play a role in success.

**Figure 6.**
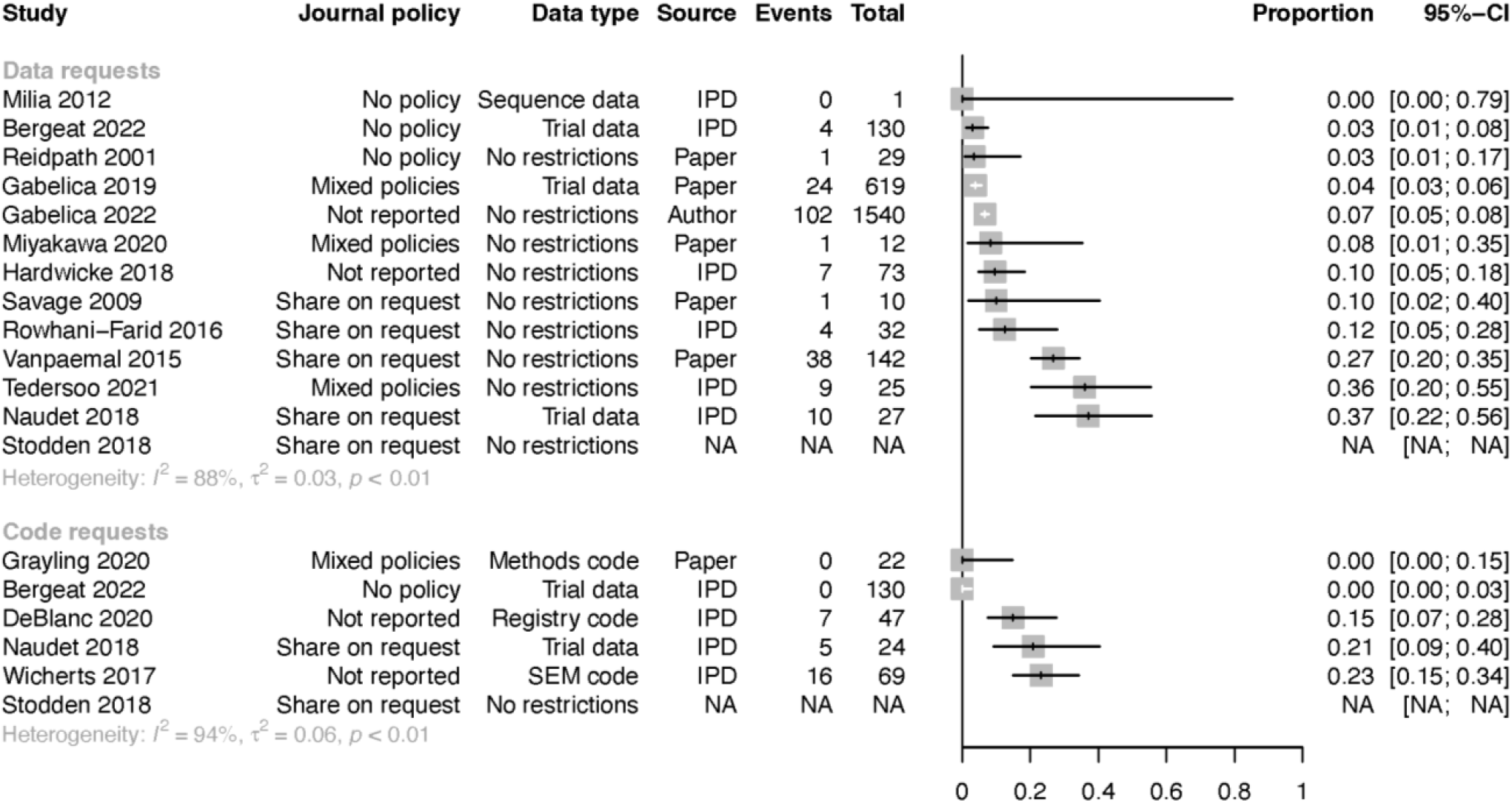
Overall success rates of private requests for data and code from published medical research. (SEM = structural equation modelling, NA = Summary data not available)

### Secondary outcomes

Insufficient data were available to evaluate the first three secondary outcome measures (i.e., outcomes concerning data and code availability statements), due to only a single study recording information about both the prevalence of statements and journal policies across a random sample of articles [15]. Similarly, very few meta-research studies recorded information on compliance with multiple data sharing policies across random samples of primary articles. This review was therefore also unable to evaluate the fourth and fifth secondary outcomes measures (i.e., direct comparison of mandatory and ‘share on request’ policies with non-mandatory data sharing policies).

However, for journals implementing mandatory data sharing policies, we estimate that 65% of primary articles (95% CI: 36-88%, 95% PI: 2-100%, k = 5, o = 28,499, I^2^ = 99%) declared data to be publicly available and 33% actually shared data (95% CI: 5-69%, k = 3, o = 429, I^2^ = 93%). In contrast, we estimate the success rate for retrieving data from authors subject to ‘share on request’ policies to be 21% (95% CI: 4-47%, k = 3, o = 166, I^2^ = 30%). For comparison, declared and actual data sharing rates under ‘encourage’ systems are estimated to be 17% (95% CI: 0-62%, k = 6, o = 1,010, I^2^ = 98%) and 8% (95% CI: 0-48%, k = 3, o = 284, I^2^ = 90%) respectively. Similarly, declared and actual sharing rates for articles published in journals with no sharing policy are estimated to be 17% (95% CI: 0-59%, k = 4, o = 686, I^2^ = 95%) and 4% (95% CI: 0-95%, k = 2, o = 198, I^2^ = 83%) respectively. Refer to Supplementary Figure 7 for the results of declared and actual public code sharing rates according to journal policies.

We were able to assess the last three secondary outcomes. Our data suggest that triallists are 31% less likely to declare data are publicly available in comparison to non-triallists (RR: 0.69, 95% CI: 0.45-1.07, 95% PI: 0.12-4.13, k = 23, I^2^ = 0%). However, when examining actual data sharing, neither group appears more or less likely to share their data than the other (RR: 0.96, 95% CI: 0.53-1.72, 95% PI: 0.15-5.95, k = 19, I^2^ = 0%) (see Figure 7). We also estimate that researchers using data derived from human participants are also 35% less likely to declare data to be publicly available than researchers working with non-human participants (RR: 0.65, 95% CI: 0.42-0.99, 95% PI: 0.12-3.61, k = 19, I^2^ = 57%). However, this decreased likelihood became more pronounced when examining actual data sharing rates (RR: 0.44, 95% CI: 0.24-0.81, 95% PI: 0.05-3.57, k = 16, I^2^ = 28%) (see Figure 8). Lastly, we estimate that researchers who declare that their data are publicly available are eight times more likely to declare code to be available also (RR: 8.03, 95% CI: 2.86-22.53, 95% PI: 0.33-194.43, k = 12, I^2^ = 32%). Additionally, researchers who are verified to have made data available are estimated to be 42 times more likely than researchers who withheld data to share code as well (RR: 42.05, 95% CI: 12.15-145.52, 95% PI: 0.94-1879.62, k = 7 I^2^ = 0%) (Supplementary Figure 8).

**Figure 7.**
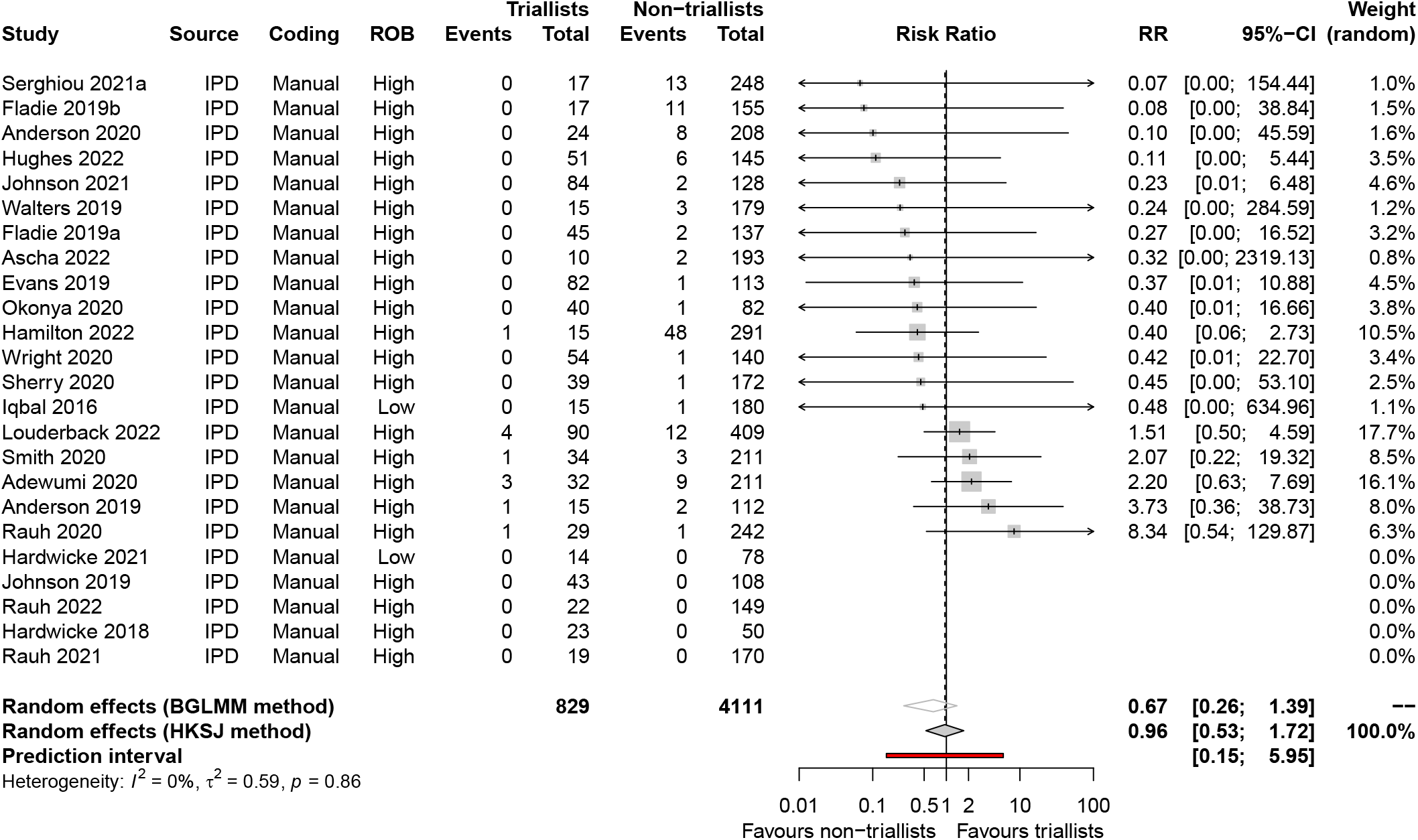
Association between trial design and data sharing (actual availability).

**Figure 8.**
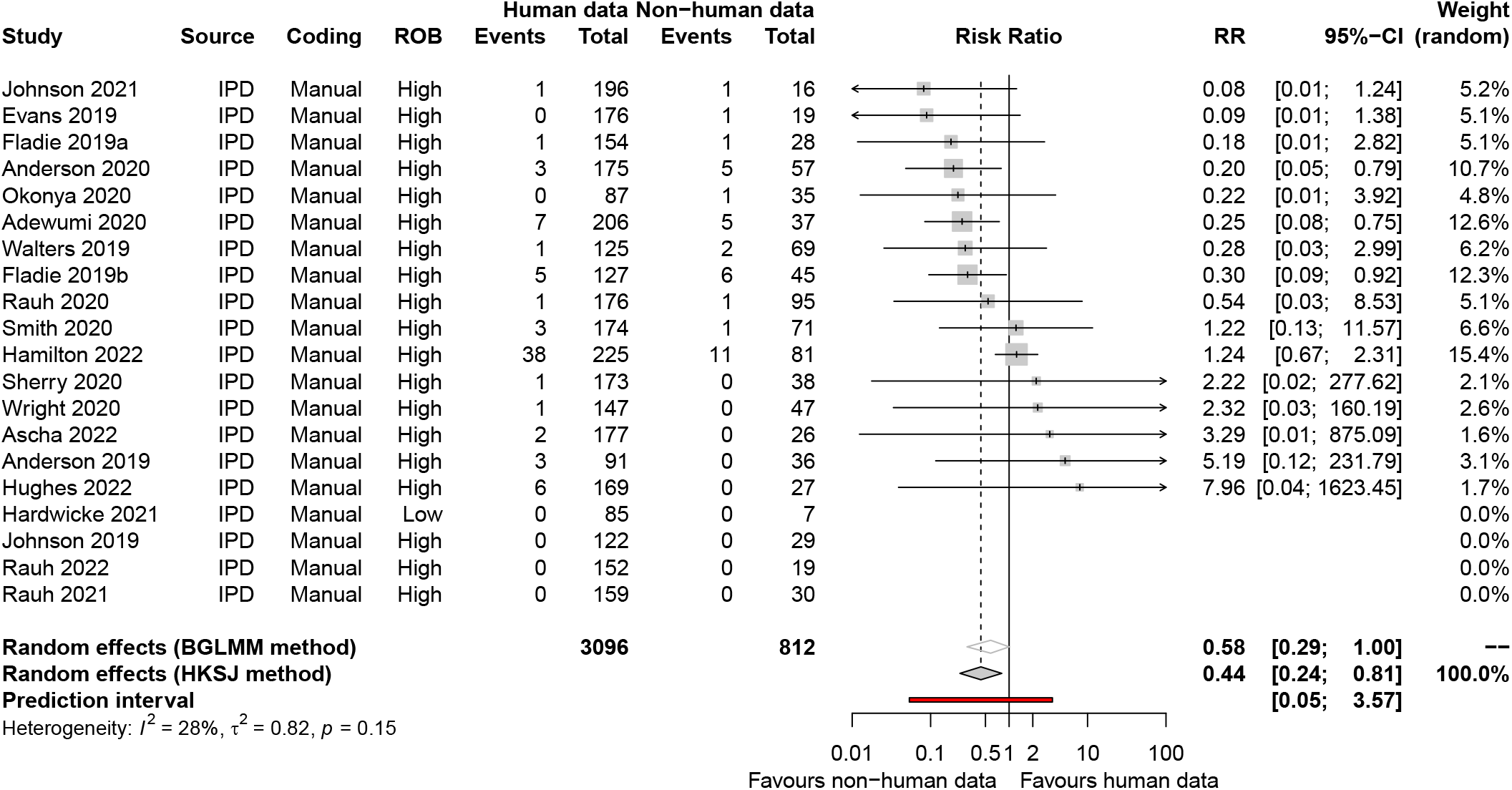
Association between research participants and data sharing (actual availability).

### Subgroup analyses

Insufficient data were available to evaluate whether prevalence estimates of public data sharing differed depending on whether primary articles were subject to any mandatory sharing policies by the funders of the research or posted as a preprint prior to publication. However, we did observe that both declared and actual public data sharing rates significantly differed according to the data type, with the highest rates of actual data sharing occurring among authors working with sequence data (57%, 95% CI: 12-96%, k = 3, o = 444, I^2^ = 86%), review data (6%, 95% CI: 0-77%, k =2, o = 372, I^2^ = 75%) then trial data (1%, 95% CI: 0-6%, k = 3, o = 235, I^2^ = 6%) (Supplementary Figures 9 & 10). Additionally, we also observed substantial differences in compliance rates with journal policies depending on the data type (Table 2). For example, estimates from a single study by Page et al [97] showed that actual data sharing rates among systematic review authors decreased from 28% for mandatory sharing policies, to 1% and 0% for encourage and no policy systems, respectively. Whereas in the context of sequence and gene expression data, decreases in actual sharing rates between mandatory policies (67% and 43%), encourage policies (57% and 43%) and no policy (46% and NA) were much less apparent.

**Table 2.**
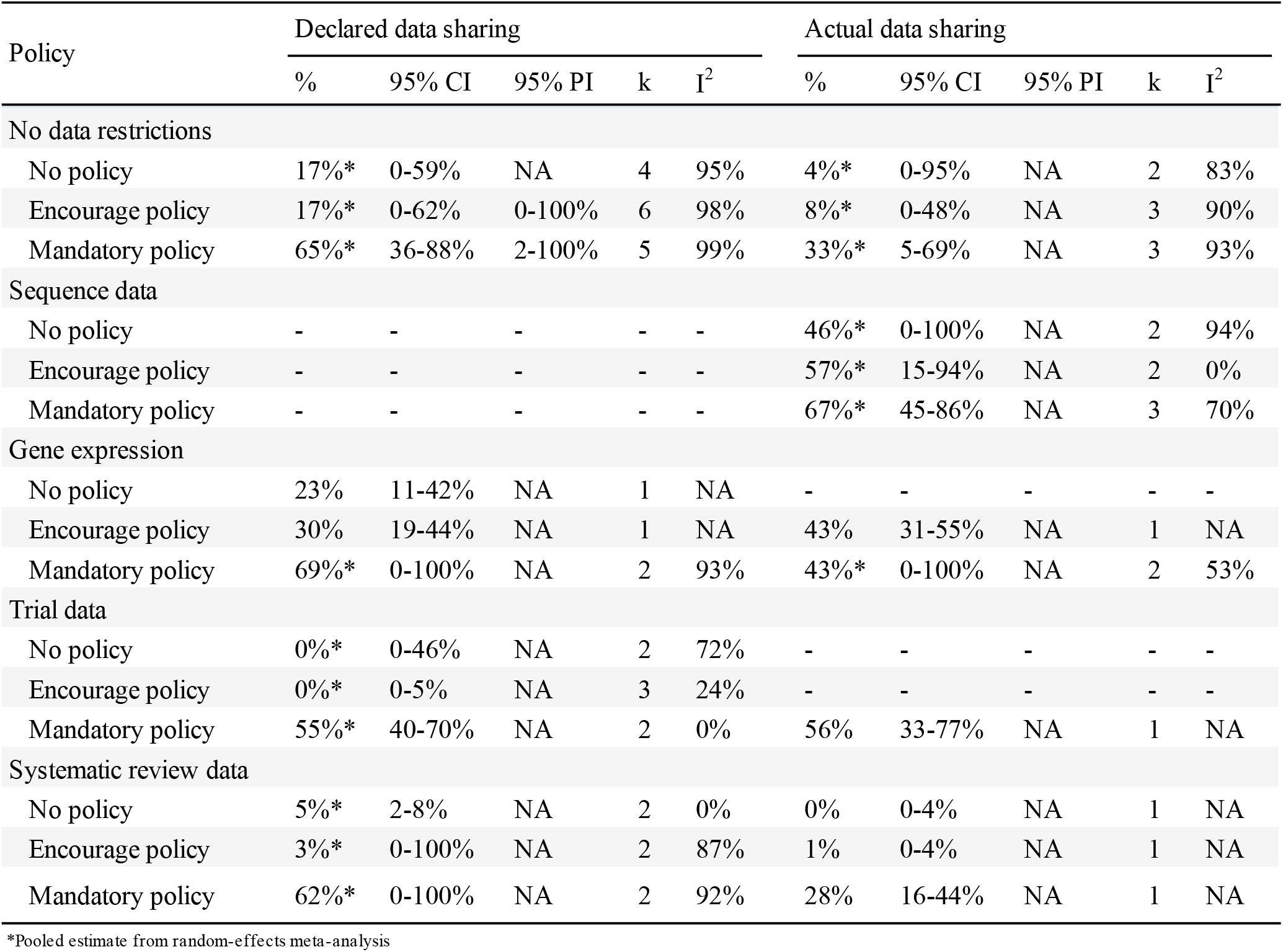
Subgroup analysis: Declared and actual public data sharing rates according to data type and journal sharing policy.

| Policy | Declared data sharing |  |  |  |  | Actual data sharing |  |  |  |  |
| --- | --- | --- | --- | --- | --- | --- | --- | --- | --- | --- |
|  | % | 95% CI | 95% PI | k | I<sup>2</sup> | % | 95% CI | 95% PI | k | I<sup>2</sup> |
| No data restrictions |  |  |  |  |  |  |  |  |  |  |
| No policy | 17%* | 0-59% | NA | 4 | 95% | 4%* | 0-95% | NA | 2 | 83% |
| Encourage policy | 17%* | 0-62% | 0-100% | 6 | 98% | 8%* | 0-48% | NA | 3 | 90% |
| Mandatory policy | 65%* | 36-88% | 2-100% | 5 | 99% | 33%* | 5-69% | NA | 3 | 93% |
| Sequence data |  |  |  |  |  |  |  |  |  |  |
| No policy | - | - | - | - | - | 46%* | 0-100% | NA | 2 | 94% |
| Encourage policy | - | - | - | - | - | 57%* | 15-94% | NA | 2 | 0% |
| Mandatory policy | - | - | - | - | - | 67%* | 45-86% | NA | 3 | 70% |
| Gene expression |  |  |  |  |  |  |  |  |  |  |
| No policy | 23% | 11-42% | NA | 1 | NA | - | - | - | - | - |
| Encourage policy | 30% | 19-44% | NA | 1 | NA | 43% | 31-55% | NA | 1 | NA |
| Mandatory policy | 69%* | 0-100% | NA | 2 | 93% | 43%* | 0-100% | NA | 2 | 53% |
| Trial data |  |  |  |  |  |  |  |  |  |  |
| No policy | 0%* | 0-46% | NA | 2 | 72% | - | - | - | - | - |
| Encourage policy | 0%* | 0-5% | NA | 3 | 24% | - | - | - | - | - |
| Mandatory policy | 55%* | 40-70% | NA | 2 | 0% | 56% | 33-77% | NA | 1 | NA |
| Systematic review data |  |  |  |  |  |  |  |  |  |  |
| No policy | 5%* | 2-8% | NA | 2 | 0% | 0% | 0-4% | NA | 1 | NA |
| Encourage policy | 3%* | 0-100% | NA | 2 | 87% | 1% | 0-4% | NA | 1 | NA |
| Mandatory policy | 62%* | 0-100% | NA | 2 | 92% | 28% | 16-44% | NA | 1 | NA |

\*Pooled estimate from random-effects meta-analysis

Finally, changes in public data and code sharing rates over time were investigated by fitting three-level mixed-effect meta-regression models to arcsine-transformed data (refer to Supplementary Table 4 for the full results). Publication year was found to be a significant moderator of declared data sharing rates (β=0.017, 95% CI: 0.008-0.025, p=0.0001, between-study I^2^ = 91%, within-study I^2^ = 9%) but not actual data sharing rates (β = 0.004, 95% CI: −0.005-0.013, p = 0.3589, between-study I^2^ = 75%, within-study I^2^ = 3%). Specifically, we note an estimated rise in declared data sharing rates from 4% in 2014 (95% CI: 2-6%, 95% PI: 0-18%) to 9% in 2020 (95% CI: 6-12%, 95% PI: 0-26%). Refer to Figure 9 for a bubble plot comparing declared data sharing rates and actual sharing rates over time. Comparatively, both declared and actual code sharing rates did not appear to have meaningfully increased over time.

**Figure 9.**
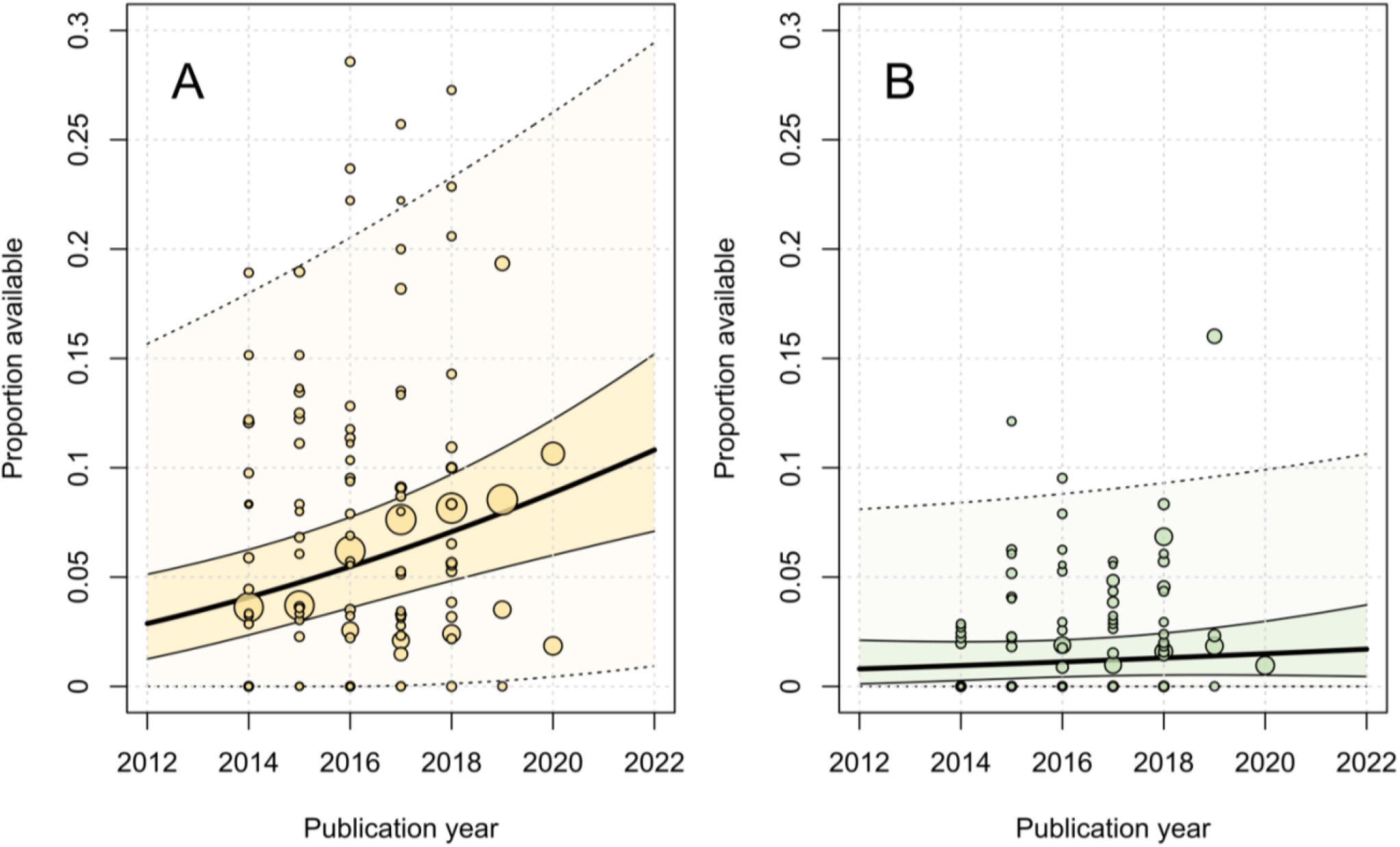
Bubble plot of declared (A) and actual (B) data sharing rates by publication year with fitted meta-regression lines, 95% confidence intervals and 95% prediction intervals. Circles are scaled relative to the natural log of the sample size.

### Sensitivity analyses

The results of the sensitivity analyses of the primary outcomes are reported in Table 3. For public data and code sharing outcomes, meta-analysis of prevalence rates using GLMMs did not result in any substantial changes to combined estimates in comparison to the standard inverse-variance aggregation methods. Similarly, limiting analyses to meta-research studies in which authors manually coded articles (i.e., removal of meta-research studies that used automated or unclear coding methods) did not result in any meaningful changes. When limiting analyses to meta-research studies where summary data were only derived from available IPD, no changes were observed to the declared data availability analysis. Insufficient data were available to evaluate whether findings from meta-research studies that assessed compliance with FAIR or were classified as low risk of bias resulted in meaningful changes to pooled estimates. Similarly, with respect to the impact of overlapping primary articles, removing the only meta-research study that was deemed to be at risk of overlapping with other included meta-research studies had no impact on any of the analyses. Lastly, we estimate declared and actual public data sharing rates for studies investigating COVID-19 (including both preprints or peer-reviewed publications) to be 9% (95% CI: 0-57%, k=3, o = 7,804, I^2^ = 95%) and 11% (95% CI: 0-76%, k=3, o = 934, I^2^ = 84%) respectively. Both of which compare favourably to our best estimates for declared (8%) and actual data sharing (2%) since 2016. The findings of the sensitivity analyses of secondary outcomes and subgroup analyses are reported in Supplementary Table 5. Most notably, we observed stronger associations between data and code sharing when including studies with no events in both groups.

**Table 3.**
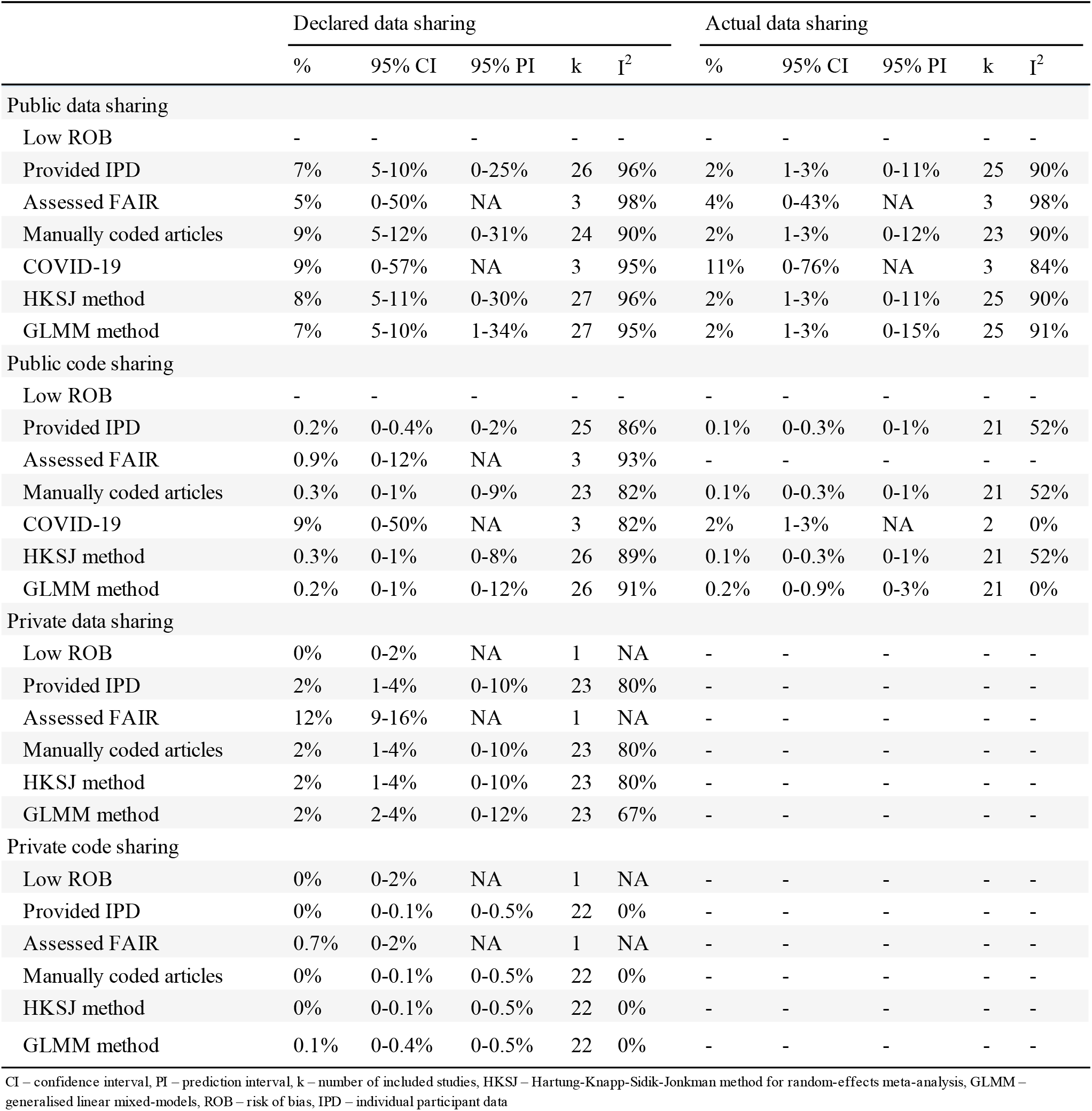
Sensitivity analyses for primary outcomes.

## DISCUSSION

### Principal findings of the review

In this, the first systematic review and IPD meta-analysis of this topic, we used multiple data sources and analytic methods to investigate public and private availability of data and code in the medical and health literature. We also examined several factors associated with sharing. Aggregation of the findings of 27 meta-research studies (which themselves examined 700,054 primary articles) suggests that on average, only 8% of medical papers published since 2016 declare that their data are publicly available. Additionally, meta-analysis of 25 meta-research studies (examining 11,873 articles), suggests that only 2% of medical papers published since 2016 will have verifiably shared their data. In comparison, estimated declared and actual code sharing rates since 2016 were even lower, with both estimated to be less than 0.5%, with little changes over time. The prediction intervals from our analyses are also relatively precise, suggesting that we now have very good estimates of data and code sharing for medical and health research since 2016.

In contrast to public availability rates, the overall success rate of privately obtaining data from authors of published medical research was observed to range between 0-37%. However, the range of success became much more variable when the scope was limited to requests made to authors who declared their data to be ‘available on request’. For private requests for code, overall success rates ranged between 0-23%, and increased to 0-43% when examining requests that were mad e to authors that declared code to be available on request. These findings are consistent with similar research conducted in fields outside of medicine [140, 143-147]. Finally, while data were not available to assess compliance with funder and institutional data sharing policies, we did observe varying compliance rates with journal data sharing policies, particularly depending on the data type.

### Review findings in context

When examining similar research conducted in other scientific fields, declared data and code sharing rates in medicine appear to be higher than some fields (e.g., Humanities, Earth Sciences and Engineering [14]), but lower than others (e.g., Experimental Biology [144, 148], Hydrology [149]). One common explanation for the differences between these sharing rates is that researchers in fields outside of the medical, health, behavioural and social sciences are more likely to make data available as they do not typically need to navigate privacy protections associated with the collection and sharing of data from human participants [150]. Our results support this notion, finding that medical researchers studying data from human participants were 66% less likely to actually make their data publicly available than those using data derived from non-human participants. This discrepancy has likely become more pronounced over time since the implementation of national and international protection laws imposing strong restrictions on the processing of personal medical data, like the US’ Health Insurance Portability and Accountability Act (HIPAA) and the European Union’s 2018 General Data Protection Regulation (GDPR) [151, 152].

We also note examples of differences in data sharing rates between medical and non-medical researchers working with the same human-derived data types. For example, in the context of human mitochondrial and Y-chromosomal data, Anagnostou and colleagues [153] observed different data sharing rates between medical genetics (64%) and forensic genetics (90%) researchers. Follow up work by the same authorship team reported that the discrepancies between sharing rates may be due to differences in cultures concerning the value of openness and transparency, as opposed to burdens associated with navigating privacy constraints [153].

### Potential implications of our findings

Our findings raise some important implications for researchers and policymakers. At a journal policymaking level, our findings suggest that overall, blanket mandatory data sharing policies do not appear to work well in medicine and health [8,146,154]. Other research suggests suboptimal compliance with mandatory sharing policies may also apply to research funders as well [155]. Furthermore, we also note such policies may vary in their effectiveness according to the data type. For example, our findings suggest that mandatory sharing policies might be an effective measure in incentivising triallists and systematic reviewers to share data but may be less effective at motivating researchers working with sequence and gene expression data to share, given the high levels of sharing under non-mandatory policies. Consequently, it may be in policymakers’ interest to periodically audit compliance with such policies, possibly triaging audits by data type, and strengthening policing if substantial non-compliance is detected. Enforcement of policies in this setting could range from simple checks of common issues (e.g., that links are present and functional [156]), up to confirming that data can be freely downloaded and are well-annotated, complete and sufficiently unprocessed.

Given average data sharing rates remain low, the medical research community could consider trialling additional incentives to increase the rate and quality of data sharing. For example, some commonly proposed strategies, beyond implementation of policies mandating sharing, include: open science badges, data embargoes, data publications, novel altmetrics, as well as changes to funding schemes to allow applicants to budget for data archival costs, and academic hiring and promotion criteria to reward sharing [87,157,158]. While such strategies have long been suggested by important medical research stakeholders such as the United States National Academy of Medicine [159], as previous research has noted, in medicine, there are more opinion pieces on the lack of incentives for researchers to share data than there have been empirical tests of these incentives. Consequently, the effectiveness of most of these strategies in medicine remain an open area of inquiry [157].

Finally, we also observed that useful data are not only difficult to retrieve from medical researchers, but also difficult to retrieve from meta-researchers who are interested in studying the very topic of data sharing; a phenomenon that we are not the first to lament [160]. As such, the results of our review also extend concerns with regards to suboptimal archival practices, beyond the medical research community to the meta-research community as well. We have also uncovered substantial amounts of research duplication among meta-research studies examining open science practices in medicine and health. Consequently, we recommend that less research attention be paid to estimating overall data and code sharing rates in medicine, particularly between 2014 and 2020.

### Strengths and limitations of the study

Our review has many methodological advantages over previous research in this area. Firstly, as data and code sharing are relatively rare events, IPD meta-analysis allowed us to bring together many imprecise findings to yield more precise estimates. Furthermore, retrieval of useful IPD from 95% of included studies has allowed us to conduct several data quality checks, identify and remove substantial amounts of redundant assessments, perform subgroup analyses not possible when conducting a meta-analysis of aggregate data, as well as minimise the risk of data availability biases. Second, the meta-analyses of our primary and secondary outcomes included more studies than the average meta-analysis of prevalence and rare events [161,162], reducing the risk of power issues, as well as making our review the largest analysis of ‘actual’ data and code sharing rates to our knowledge. Similarly, we had more than double the recommended number of estimates per covariate for our meta-regression analyses, minimising the risk of issues such as overfitting [163]. Third, the review included robustness checks using generalised linear mixed models, which have been recommended over conventional meta-analyses of arcsine-transformed proportions [164]. The use of GLMMs also allowed for analysis of studies with zero events in both groups which can alter conclusions in some circumstances [165], as well as circumvent the need to add arbitrary continuity corrections when meta-analysing risk ratios which can bias results [27].

Nevertheless, our review was not without limitations. First, we may have missed relevant literature due to challenges in designing the search strategies (e.g., lack of controlled vocabulary, variations in the way meta-research studies described themselves) and limiting searches to predominantly English-language databases. Second, we were unable to include the findings of nine studies due to the inability to source IPD or useable summary data. However, given 97% of primary articles examined by the excluded studies were at high risk of overlap with studies that were included in the analysis, we do not think their omission would have substantially altered our findings. Third, only one author performed IPD checks and harmonisation. Fourth, we assumed that authors will always declare in-text when data and/or code have been made publicly available, which previous studies have shown is not always the case [92]. However, this appears to be an uncommon practice, and therefore was unlikely to have significantly impacted our results. Finally, despite efforts to ensure studies were clinically homogeneous, our meta-analyses of proportions demonstrated high levels of statistical heterogeneity. However, given three-quarters of published meta-analyses of proportions report I^2^ values greater than 90% [161], the statistic’s usefulness for assessing heterogeneity in this context is debated. Consequently, evidence synthesis researchers have recommended greater priority be placed on visually inspecting forest plots and prediction interval widths instead [161]. Therefore, while we acknowledge these high I^2^ values, given the consistency of study methods and reported estimates (refer to Figure 5 for an illustrative example), we do not believe these values indicate concerning levels of variability in this context.

### Unanswered questions and future research

We note several questions that have not been answered by our review. Our review was unable to comment on compliance rates with mandatory sharing policies introduced by medical research institutions and funders as we could not find relevant meta-research on this question. Furthermore, while we were able to explore how sharing rates differed according to data type, our analyses were restricted to a limited number of studies effectively examining four types of data (trial data, systematic review data, gene expression data and sequence data). Consequently, we were unable to establish precise estimates for compliance rates, nor comment on sharing rates for the myriad of other types of data that medical researchers have discussed with respect to data sharing. For example, model data [166], imaging data [167], flow cytometry data [168], spectroscopic data [169], diffraction data [170,171], and qualitative data [172] to name a few. Both are areas worthy of future empirical meta-research.

With regards to future research, we hope that the data that we have collected and harmonised for this review will serve as a useful resource to track changes in data and code sharing in medicine beyond 2020, as well as explore other factors that we were unable to assess (e.g., association between preprinting practices and sharing) or had not considered (e.g., association between career stage and sharing [173]). However, as we have been able to establish precise estimates of public data and code sharing rates, we do not think additional research that examines high-level data and code sharing rates in medicine between 2014-2020 is warranted.

## CONCLUSION

The results of the current review suggest that while increasing numbers of medical and health researchers are stating that their data are publicly available, declaration rates remain uncommon, and not all declarations lead to the stated data. In contrast, code sharing rates remain persistently low across medicine. We also note large variability in success rates in privately obtaining data and code from authors of published medical research. While no data were available to evaluate the effectiveness of funder and institutional policies on data sharing, assessments of journal policies suggest that mandatory sharing policies are more effective than non-mandatory policies, as well as may demonstrate varying success rates according to the data type -a finding that may be informative for policymakers when designing policies and allocating resources to audit compliance.

## Data Availability

Summary level data and the code required to reproduce all the findings of the review are freely available on the Open Science Framework (DOI: 10.17605/OSF.IO/U3YRP) under a Creative Commons Zero v1.0 Universal (CC0 1.0) license. Harmonised versions of IPD that were originally made publicly available can be shared on request, whereas to preserve the rights of data owners, harmonised versions of IPD that were shared privately with the review team will only be released with the permission of the data guarantor of the relevant meta-research study. To request harmonised IPD please follow the instructions on the project's Open Science Framework page (https://osf.io/stnk3).

https://www.doi.org/10.17605/OSF.IO/U3YRP

## Disclosures

### Ethics approval

Not applicable.

### Availability of data and materials

Summary level data and the code required to reproduce all the findings of the review are freely available on the Open Science Framework (DOI: 10.17605/OSF.IO/U3YRP) under a Creative Commons Zero v1.0 Universal (CC0 1.0) license. Harmonised versions of IPD that was originally made publicly available can be shared on request, whereas to preserve the rights of data owners, harmonised versions of IPD that were shared privately with the review team will only be released with the permission of the data guarantor of the relevant meta-research study. To request harmonised IPD please follow the instructions on the project’s Open Science Framework page (https://osf.io/stnk3).

### Competing interests

The authors declare that they have no competing interests

## Funding

No funding was received for this study. DGH is a PhD candidate supported by an Australian Commonwealth Government Research Training Program Scholarship. MJP is supported by an Australian Research Council Discovery Early Career Researcher Award (DE200101618).

## Acknowledgements

We thank Steve McDonald for his assistance with the literature searches, A/Prof Sue Finch for her advice on the statistical analyses and all the meta-researchers who took the time to prepare data and address clarifications for the review.

## Notes

### Competing Interest Statement

The authors have declared no competing interest.

### Clinical Protocols

https://doi.org/10.12688/f1000research.53874.2

## References

1. Shahin MH, Bhattacharya S, Silva D, Kim S, Burton J, Podichetty J, et al. Open Data Revolution in Clinical Research: Opportunities and Challenges. Clinical and Translational Science. 2020;13(4):665–74.

2. Kim J, Kim S, Cho HM, Chang JH, Kim SY. Data sharing policies of journals in life, health, and physical sciences indexed in Journal Citation Reports. PeerJ. 2020 Oct 13;8:e9924.

3. Hamilton DG, Fraser H, Hoekstra R, Fidler F. Journal policies and editors’ opinions on peer review. eLife. 2020 Nov 19;9:e62529.

4. Resnik DB, Morales M, Landrum R, Shi M, Minnier J, Vasilevsky NA, et al. Effect of impact factor and discipline on journal data sharing policies. Accountability in Research. 2019 Apr 3;26(3):139–56.

5. DeVito NJ, French L, Goldacre B. Noncommercial Funders’ Policies on Trial Registration, Access to Summary Results, and Individual Patient Data Availability. JAMA. 2018 Apr 24;319(16):1721–3.

6. Gaba JF, Siebert M, Dupuy A, Moher D, Naudet F. Funders’ data-sharing policies in therapeutic research: A survey of commercial and non-commercial funders. PLOS ONE. 2020 Aug 20;15(8):e0237464.

7. The White House. OSTP Issues Guidance to Make Federally Funded Research Freely Available Without Delay. August 25th, 2022. https://www.whitehouse.gov/wp-content/uploads/2022/08/08-2022-OSTP-Public-Access-Memo.pdf

8. Milia N, Congiu A, Anagnostou P, Montinaro F, Capocasa M, Sanna E, Bisol GD. Mine, Yours, Ours? Sharing Data on Human Genetic Variation. PLoS ONE. 2012; 7: e37552. DOI: 10.1371/journal.pone.0037552.

9. Rufiange M, Rousseau-Blass F, Pang DSJ. Incomplete reporting of experimental studies and items associated with risk of bias in veterinary research. Veterinary Record Open. 6;2019. DOI: 10.1136/vetreco-2018-000322.

10. Witwer KW. Data Submission and Quality in Microarray-Based MicroRNA Profiling. Clinical Chemistry. 2013; 59: 392–400. DOI: 10.1373/clinchem.2012.193813.

11. Zavalis EA, Ioannidis JPA. A metaepidemiological assessment of transparency indicators of infectious disease models. 2022; DOI: 10.1101/2022.04.11.22273744.

12. Gabelica M, Cavar J, Puljak L. Authors of trials from high-ranking anesthesiology journals were not willing to share raw data. Journal of Clinical Epidemiology. 2019;109: 111–116. DOI: 10.1016/j.jclinepi.2019.01.012.

13. Nguyen PY, Kanukula R, McKenzie JE, Alqaidoom Z, Brennan SE, Haddaway NR, et al. Changing patterns in reporting and sharing of review data in systematic reviews with metaanalysis of the effects of interventions: cross sectional meta-research study. BMJ. 2022 Nov 22; 379: e072428. DOI: 10.1136/bmj-2022-072428

14. Serghiou S, Contopoulos-Ioannidis DG, Boyack KW, Riedel N, Wallach JD, Ioannidis JPA. Assessment of transparency indicators across the biomedical literature: How open is open? PLOS Biology. 2021; 19: e3001107. DOI: 10.1371/journal.pbio.3001107.

15. Hamilton DG, Page MJ, Finch S, Everitt S, Fidler F. How often do cancer researchers make their data and code available and what factors are associated with sharing? BMC Medicine. 2022; 20. DOI: 10.1186/s12916-022-02644-2.

16. Hamilton DG, Fraser H, Fidler F, Rowhani-Farid A, Hong K, Page MJ. Rates and predictors of data and code sharing in the medical and health sciences: A systematic review and individual participant data meta-analysis. Open Science Framework, May 28, 2021. DOI: 10.17605/OSF.IO/7SX8U.

17. Hamilton DG, Fraser H, Fidler F, McDonald S, Rowhani-Farid A, Hong K & Page MJ. Rates and predictors of data and code sharing in the medical and health sciences: Protocol for a systematic review and individual participant data metaanalysis. [version 2; peer review: 2 approved]. F1000Research 2021, 10: 491. DOI: 10.12688/f1000research.53874.2.

18. Page MJ, McKenzie JE, Bossuyt PM, et al.: The PRISMA 2020 statement: an updated guideline for reporting systematic reviews. Syst Rev. 2021; 10(1): 89.

19. Stewart LA, Clarke M, Rovers M, et al.: Preferred Reporting Items for Systematic Review and Meta-Analyses of individual participant data: the PRISMA-IPD Statement. JAMA. 2015; 313(16): 1657–1665.

20. Hamilton DG, Fraser H, Fidler F, McDonald S, Rowhani-Farid A, Hong K & Page MJ. A review of data and code sharing rates in medical and health research. Open Science Framework, 2023. https://doi.org/10.17605/OSF.IO/H75V4.

21. Haddaway NR, Grainger MJ, Gray, CT. citationchaser: An R package and Shiny app for forward and backward citations chasing in academic searching. 2021. DOI: 10.5281/zenodo.4543513.

22. Hansen C, Bero L, Hróbjartsson A, et al.: Conflicts of interest and recommendations in clinical guidelines, opinion pieces, and narrative reviews. Cochrane Database Syst Rev. 2019; 10.

23. Page MJ, McKenzie JE, Kirkham J, Dwan K, Kramer S, Green S, Forbes A. Bias due to selective inclusion and reporting of outcomes and analyses in systematic reviews of randomised trials of healthcare interventions. Cochrane Database Syst Rev. 2014; 2014(10): MR000035.

24. Porter SJ & Hook DW. Recategorising research: Mapping from FoR 2008 to FoR 2020 in Dimensions. arXiv. 2022: 2209.00104v1. DOI: 10.48550/arXiv.2209.00104.

25. Wilkinson M, Dumontier M, Aalbersberg I, et al. The FAIR Guiding Principles for scientific data management and stewardship. Sci Data. 2016; 3: 160018.

26. IntHout J, Ioannidis JP, Borm GF. Tthe Hartung-Knapp-Sidik-Jonkman method for random effects meta-analysis is straightforward and considerably outperforms the standard DerSimonian-Laird method. BMC Med Res Methodol 2014; 14(1): 25. DOI: 10.1186/1471-2288-14-25.

27. Sweeting MJ, Sutton A, Lambert PC. What to add to nothing? Use and avoidance of continuity corrections in meta-analysis of sparse data. Statistics in Medicine. 2004; 23(9): 1351–75.

28. Balduzzi S, Rücker G, Schwarzer G. How to perform a meta-analysis with R: a practical tutorial, Evidence-Based Mental Health 2019; 22: 153–160. DOI: 10.1136/ebmental-2019-300117.

29. Viechtbauer, W. Conducting meta-analyses in R with the metafor package. Journal of Statistical Software, 2010; 36(3): 1–48. DOI: 10.18637/jss.v036.i03.

30. Lin L, Chu H (2022). altmeta: Alternative Meta-Analysis Methods. R package version 4.1. https://CRAN.R-project.org/package=altmeta.

31. McGuinness, LA, Higgins, JPT. Risk-of-bias VISualization (robvis): An R package and Shiny web app for visualizing risk-of-bias assessments. Res Syn Meth. 2020;1-7. https://doi.org/10.1002/jrsm.1411

32. Stijnen T, Hamza TH, Özdemir P. Random effects meta-analysis of event outcome in the framework of the generalized linear mixed model with applications in sparse data. Statist Med. 2010 Dec 20;29(29):3046–67.

33. Chu H, Nie L, Chen Y, Huang Y, Sun W. Bivariate random effects models for metaanalysis of comparative studies with binary outcomes: methods for the absolute risk difference and relative risk. Statistical Methods in Medical Research 2012; 21(6):621–633. DOI: 10.1177/0962280210393712.

34. Adewumi MT, Vo N, Tritz D, Beaman J, Vassar M. An evaluation of the practice of transparency and reproducibility in addiction medicine literature. Addictive Behaviors. 2021; 112: 106560. DOI: 10.1016/j.addbeh.2020.106560.

35. Alsheikh-Ali AA, Qureshi W, Al-Mallah MH, Ioannidis JPA. Public Availability of Published Research Data in High-Impact Journals. PLoS ONE. 2011; 6: e24357. DOI: 10.1371/journal.pone.0024357.

36. Anderson JM, Niemann A, Johnson AL, Cook C, Tritz D, Vassar M. Transparent, Reproducible, and Open Science Practices of Published Literature in Dermatology Journals: Cross-Sectional Analysis. JMIR Dermatology. 2019; 2: e16078. DOI: 10.2196/16078.

37. Anderson JM, Wright B, Rauh S, Tritz D, Horn J, Parker I, Bergeron D, Cook S, Vassar M. Evaluation of indicators supporting reproducibility and transparency within cardiology literature. Heart. 2020; 107: 120–126. DOI: 10.1136/heartjnl-2020-316519.

38. Ascha M, Katabi L, Stevens E, Gatherwright J, Vassar M. Reproducible Research Practices in the Plastic Surgery Literature. Plastic & Reconstructive Surgery. 2022; 149: 810e–823e. DOI: 10.1097/prs.0000000000008956.

39. Bergeat D, Lombard N, Gasmi A, Le Floch B, Naudet F. Data Sharing and Reanalyses Among Randomized Clinical Trials Published in Surgical Journals Before and After Adoption of a Data Availability and Reproducibility Policy. JAMA Network Open. 2022; 5: e2215209. DOI: 10.1001/jamanetworkopen.2022.15209.

40. Bonetti AF, Tonin FS, Lucchetta RC, Pontarolo R, Fernandez-Llimos F. Methodological standards for conducting and reporting metaanalyses: Ensuring the replicability of metaanalyses of pharmacist-led medication review. Research in Social and Administrative Pharmacy. 2022; 18: 2259–2268. DOI: 10.1016/j.sapharm.2021.06.002.

41. Borghi JA, Payne C, Ren L, Woodward AL, Wong C, Stave C. Open Science and COVID-19 Randomized Controlled Trials: Examining Open Access, Preprinting, and Data Sharing-Related Practices During the Pandemic. 2022; DOI: 10.1101/2022.08.10.22278643.

42. Cenci MS, Franco MC, Raggio DP, Moher D, Pereira-Cenci T. Transparency in clinical trials: Adding value to paediatric dental research. International Journal of Paediatric Dentistry. 2020; 31: 4–13. DOI: 10.1111/ipd.12769.

43. Colavizza G, Hrynaszkiewicz I, Staden I, Whitaker K, McGillivray B. The citation advantage of linking publications to research data. PLOS ONE. 2020; 15: e0230416. DOI: 10.1371/journal.pone.0230416.

44. Collins A, Alexander R. Reproducibility of COVID-19 preprints. Scientometrics. 2022; 127: 4655–4673. DOI: 10.1007/s11192-022-04418-2.

45. Danchev V, Min Y, Borghi J, Baiocchi M, Ioannidis JPA. Evaluation of Data Sharing After Implementation of the International Committee of Medical Journal Editors Data Sharing Statement Requirement. JAMA Network Open. 2021; 4: e2033972. DOI: 10.1001/jamanetworkopen.2020.33972.

46. DeBlanc J, Kay B, Lehrich J, Kamdar N, Valley TS, Ayanian JZ, Nallamothu BK. Availability of Statistical Code From Studies Using Medicare Data in General Medical Journals. JAMA Internal Medicine. 2020; 180: 905. DOI: 10.1001/jamainternmed.2020.0671.

47. Duan Y, Luo J, Zhao L, Zhang X, Miao J, Moher D, Bian Z. Reporting and data sharing level for COVID-19 vaccine trials: A cross-sectional study. eBioMedicine. 2022; 78: 103962. DOI: 10.1016/j.ebiom.2022.103962.

48. Errington TM, Denis A, Perfito N, Iorns E, Nosek BA. Challenges for assessing replicability in preclinical cancer biology. eLife. 2021;10. DOI: 10.7554/elife.67995.

49. Evans S, Fladie IA, Anderson JM, Tritz D, Vassar M. Evaluation of Reproducible and Transparent Research Practices in Sports Medicine Research: A Cross-sectional study. 2019; DOI: 10.1101/773473.

50. Federer LM, Belter CW, Joubert DJ, Livinski A, Lu Y-L, Snyders LN, Thompson H. Data sharing in PLOS ONE: An analysis of Data Availability Statements. PLOS ONE. 2018; 13: e0194768. DOI: 10.1371/journal.pone.0194768.

51. Fladie IA, Adewumi TM, Vo NH, Tritz DJ, Vassar MB. Atn Evaluation of Nephrology Literature for Transparency and Reproducibility Indicators: Cross-Sectional Review. Kidney International Reports. 2020; 5: 173–181. DOI: 10.1016/j.ekir.2019.11.001.

52. Fladie IA, Evans S, Checketts J, Tritz D, Norris B, Vassar M. Can Orthopaedics become the Gold Standard for Reproducibility? A Roadmap to Success. 2019; DOI: 10.1101/715144.

53. Gabelica M, Bojcic R, Puljak L. Many researchers were not compliant with their published data sharing statement: a mixedmethods study. Journal of Clinical Epidemiology. 2022; 150: 33–41. DOI: 10.1016/j.jclinepi.2022.05.019.

54. Gkiouras K, Nigdelis MP, Grammatikopoulou MG, Goulis DG. Tracing open data in emergencies: The case of the COVID-19 pandemic. European Journal of Clinical Investigation. 2020;50. DOI: 10.1111/eci.13323.

55. Gorman DM. Availability of Research Data in High-Impact Addiction Journals with Data Sharing Policies. Science and Engineering Ethics. 2020; 26: 1625–1632. DOI: 10.1007/s11948-02000203-7.

56. Grant R, Hrynaszkiewicz I. The impact on authors and editors of introducing Data Availability Statements at Nature journals. International Journal of Digital Curation. 2018; 13: 195–203. DOI: 10.2218/ijdc.v13i1.614.

57. Grayling MJ, Wheeler GM. A review of available software for adaptive clinical trial design. Clinical Trials. 2020; 17: 323–331. DOI: 10.1177/1740774520906398.

58. Hanson KA, Almeida N, Traylor JI, Rajagopalan D, Johnson J. Profile of Data Sharing in the Clinical Neurosciences. Cureus. 2020; DOI: 10.7759/cureus.9927.

59. Hardwicke TE, Ioannidis JPA. Populating the Data Ark: An attempt to retrieve, preserve, and liberate data from the most highly-cited psychology and psychiatry articles. PLOS ONE. 2018; 13: e0201856. DOI: 10.1371/journal.pone.0201856.

60. Hardwicke TE, Thibault RT, Kosie JE, Wallach JD, Kidwell MC, Ioannidis JPA. Estimating the Prevalence of Transparency and Reproducibility-Related Research Practices in Psychology (2014-2017). Perspectives on Psychological Science. 2021; 17: 239–251. DOI: 10.1177/1745691620979806.

61. Heckerman G, Tzng E, Campos-Melendez A, Ekwueme C, Mueller AL. Accessibility and Reproducible Research Practices in Cardiovascular Literature. 2022; DOI: 10.1101/2022.07.06.498942.

62. Heller N, Rickman J, Weight CJ, Papanikolopoulos N. The Role of Publicly Available Data in MICCAI Papers from 2014 to 2018. arXiv 2019; 1908.06830. DOI: 10.48550/arXiv.1908.06830.

63. Helliwell JA, Shelton B, Mahmood H, Blanco-Colino R, Fitzgerald JE, Harrison EM, Bhangu A, Chapman SJ. Transparency in surgical randomized clinical trials: cross-sectional observational study. BJS Open. 2020; 4: 977–984. DOI: 10.1002/bjs5.50333.

64. Huang Y-N, Patel NA, Mehta JH, Ginjala S, Brodin P, Gray CM, Patel YM, Cowell LG, Burkhardt AM, Mangul S. Data Availability of Open T-Cell Receptor Repertoire Data, a Systematic Assessment. Frontiers in Systems Biology. 2022; 2. DOI: 10.3389/fsysb.2022.918792.

65. Hughes T, Niemann A, Tritz D, Boyer K, Robbins H, Vassar M. Transparent and Reproducible Research Practices in the Surgical Literature. bioRxiv 2019; 779702. DOI: 10.1101/779702v1

66. Ioannidis JPA, Allison DB, Ball CA, Coulibaly I, Cui X, Culhane AC, Falchi M, Furlanello C, Game L, Jurman G, et al. Repeatability of published microarray gene expression analyses. Nature Genetics. 2009; 41: 149–155. DOI: 10.1038/ng.295.

67. Ioannidis JPA, Polyzos NP, Trikalinos TA. Selective discussion and transparency in microarray research findings for cancer outcomes. European Journal of Cancer. 2007; 43: 1999–2010. DOI: 10.1016/j.ejca.2007.05.019.

68. Iqbal SA, Wallach JD, Khoury MJ, Schully SD, Ioannidis JPA. Reproducible Research Practices and Transparency across the Biomedical Literature. PLOS Biology. 2016; 14: e1002333. DOI: 10.1371/journal.pbio.1002333.

69. Jalali MS, DiGennaro C, Sridhar D. Transparency Assessment of COVID-19 Models. The Lancet Global Health, 2020; 8(12): e1459–e1460. DOI: 10.1101/2020.07.18.20156851.

70. Janssen MA, Pritchard C, Lee A. On code sharing and model documentation of published individual and agent-based models. Environmental Modelling & Software. 2020; 134: 104873. DOI: 10.1016/j.envsoft.2020.104873.

71. Johnson AL, Torgerson T, Skinner M, Hamilton T, Tritz D, Vassar M. An assessment of transparency and reproducibility-related research practices in otolaryngology. The Laryngoscope. 2019; 130: 1894–1901. DOI: 10.1002/lary.28322.

72. Johnson B, Rauh S, Tritz D, Schiesel M, Vassar M. Evaluating Reproducibility and Transparency in Emergency Medicine Publications. Western Journal of Emergency Medicine. 2021; 22: 963–971. DOI: 10.5811/westjem.2021.3.50078.

73. Johnson JN, Hanson KA, Jones CA, Grandhi R, Guerrero J, Rodriguez J. Data Sharing in Neurosurgery and Neurology Journals. Cureus. 2018; DOI: 10.7759/cureus.2680.

74. Jurburg SD, Konzack M, Eisenhauer N, Heintz-Buschart A. The archives are half-empty: an assessment of the availability of microbial community sequencing data. Communications Biology. 2020;3. DOI: 10.1038/s42003-020-01204-9.

75. Kaufmann I, Gattrell WT. Assessment of journal compliance with data sharing guidelines from the International Committee of Medical Journal Editors (ICMJE). Current Medical Research and Opinion. 2019 Apr 11; 35(sup2): 31–41. DOI: 10.1080/03007995.2019.1583496

76. Kemper JM, Rolnik DL, Mol BWJ, Ioannidis JPA. Reproducible research practices and transparency in reproductive endocrinology and infertility articles. Fertility and Sterility. 2020; 114: 1322–1329. DOI: 10.1016/j.fertnstert.2020.05.020.

77. Kirouac DC, Cicali B, Schmidt S. Reproducibility of Quantitative Systems Pharmacology Models: Current Challenges and Future Opportunities. CPT: Pharmacometrics & Systems Pharmacology. 2019; 8: 205–210. DOI: 10.1002/psp4.12390.

78. Kobres P-Y, Chretien J-P, Johansson MA, Morgan JJ, Whung P-Y, Mukundan H, Del Valle SY, Forshey BM, Quandelacy TM, Biggerstaff M, et al. A systematic review and evaluation of Zika virus forecasting and prediction research during a public health emergency of international concern. PLOS Neglected Tropical Diseases. 2019; 13: e0007451. DOI: 10.1371/journal.pntd.0007451.

79. Littmann M, Selig K, Cohen-Lavi L, Frank Y, Honigschmid P, Kataka E, Mosch A, Qian K, Ron A, Schmid S, et al. Validity of machine learning in biology and medicine increased through collaborations across fields of expertise. Nature Machine Intelligence. 2020; 2: 18–24. DOI: 10.1038/s42256-019-0139-8.

80. Lopez-Nicolas R, Lopez-Lopez JA, Rubio-Aparicio M, Sanchez-Meca J. A meta-review of transparency and reproducibility-related reporting practices in published meta-analyses on clinical psychological interventions (2000-2020). Behavior Research Methods. 2021; 54: 334–349. DOI: 10.3758/s13428-021-01644-z.

81. Louderback ER, Gainsbury SM, Heirene RM, Amichia K, Grossman A, Bernhard BJ, LaPlante DA. Open Science Practices in Gambling Research Publications (2016-2019): A Scoping Review. Journal of Gambling Studies. 2022; DOI: 10.1007/s10899-022-10120-y.

82. McGuinness LA, Sheppard AL. A descriptive analysis of the data availability statements accompanying medRxiv preprints and a comparison with their published counterparts. PLOS ONE. 2021; 16: e0250887. DOI: 10.1371/journal.pone.0250887.

83. Meyer A, Faverjon C, Hostens M, Stegeman A, Cameron A. Systematic review of the status of veterinary epidemiological research in two species regarding the FAIR guiding principles. BMC Veterinary Research. 2021;17. DOI: 10.1186/s12917-021-02971-1.

84. Miyakawa T. No raw data, no science: another possible source of the reproducibility crisis. Molecular Brain. 2020; 13. DOI: 10.1186/s13041-020-0552-2.

85. Munkholm K, Faurholt-Jepsen M, Ioannidis JPA, Hemkens LG. Consideration of confounding was suboptimal in the reporting of observational studies in psychiatry: a meta-epidemiological study. Journal of Clinical Epidemiology. 2020; 119: 75–84. DOI: 10.1016/j.jclinepi.2019.12.002.

86. Munro BA, Bergen P, Pang DSJ. Randomization, blinding, data handling and sample size estimation in papers published in Veterinary Anaesthesia and Analgesia in 2009 and 2019. Veterinary Anaesthesia and Analgesia. 2022; 49: 18–25. DOI: 10.1016/j.vaa.2021.09.004.

87. Naudet F, Sakarovitch C, Janiaud P, Cristea I, Fanelli D, Moher D, Ioannidis JPA. Data sharing and reanalysis of randomized controlled trials in leading biomedical journals with a full data sharing policy: survey of studies published in The BMJ and PLOS Medicine. BMJ. 2018;k400. DOI: 10.1136/bmj.k400.

88. Noor MAF, Zimmerman KJ, Teeter KC. Data Sharing: How Much Doesn’t Get Submitted to GenBank? PLoS Biology. 2006; 4: e228. DOI: 10.1371/journal.pbio.0040228.

89. Norris E, He Y, Loh R, West R, Michie S. Assessing Markers of Reproducibility and Transparency in Smoking Behaviour Change Intervention Evaluations. Journal of Smoking Cessation. 2021; 2021: 1–12. DOI: 10.1155/2021/6694386.

90. Norris E, Sulevani I, Finnerty AN, Castro O. Assessing Open Science practices in physical activity behaviour change intervention evaluations. BMJ Open Sport & Exercise Medicine. 2022; 8: e001282. DOI: 10.1136/bmjsem-2021-001282.

91. Ntzani EE, Ioannidis JP. Predictive ability of DNA microarrays for cancer outcomes and correlates: an empirical assessment. The Lancet. 2003 Nov; 362(9394):1439–44. DOI: 10.1016/S0140-6736(03)14686-7.

92. Nuijten MB, Borghuis J, Veldkamp CLS, Dominguez-Alvarez L, van Assen MALM250 cl:115, Wicherts JM. Journal Data Sharing Policies and Statistical Reporting Inconsistencies in Psychology. Collabra: Psychology. 2017; 3. DOI: 10.1525/collabra.102.

93. Nutu D, Gentili C, Naudet F, Cristea IA. Open science practices in clinical psychology journals: An audit study. Journal of Abnormal Psychology. 2019; 128: 510–516. DOI: 10.1037/abn0000414.

94. Ochsner SA, Steffen DL, Stoeckert CJ, McKenna NJ. Much room for improvement in deposition rates of expression microarray datasets. Nat Methods. 2008 Dec; 5(12): 991–991.

95. Okonya O, Rorah D, Tritz D, Umberham BA, Wiley M, Vassar M. Analysis of Practices to Promote Reproducibility and Transparency in Anaesthesiology Research: Are Important Aspects “Hidden Behind the Drapes?” bioRxiv 2019; 729129. DOI: 10.1101/729129v1

96. Page MJ, Altman DG, Shamseer L, McKenzie JE, Ahmadzai N, Wolfe D, Yazdi F, Catal√°-L√≥pez F, Tricco AC, Moher D. Reproducible research practices are underused in systematic reviews of biomedical interventions. Journal of Clinical Epidemiology. 2018; 94: 8–18. DOI: 10.1016/j.jclinepi.2017.10.017.

97. Page MJ, Nguyen P-Y, Hamilton DG, Haddaway NR, Kanukula R, Moher D, McKenzie JE. Data and code availability statements in systematic reviews of interventions were often missing or inaccurate: a content analysis. Journal of Clinical Epidemiology. 2022; 147: 1–10. DOI: 10.1016/j.jclinepi.2022.03.003.

98. Papageorgiou SN, Antonoglou GN, Martin C, Eliades T. Methods, transparency and reporting of clinical trials in orthodontics and periodontics. Journal of Orthodontics. 2019; 46: 101–109. DOI: 10.1177/1465312519842315.

99. Park HY, Suh CH, Woo S, Kim PH, Kim KW. Quality Reporting of Systematic Review and Meta-Analysis According to PRISMA 2020 Guidelines: Results from Recently Published Papers in the Korean Journal of Radiology. Korean Journal of Radiology. 2022; 23: 355. DOI: 10.3348/kjr.2021.0808.

100. Pellen C, Caquelin L, Jouvance-Le Bail A, Gaba J, Verin M, Moher D, Ioannidis JPA, Naudet F. Intent to share Annals of Internal Medicine’s trial data was not associated with data re-use. Journal of Clinical Epidemiology. 2021; 137: 241–249. DOI: 10.1016/j.jclinepi.2021.04.011.

101. Piwowar HA, Chapman WW. Public sharing of research datasets: A pilot study of associations. Journal of Informetrics. 2010; 4: 148–156. DOI: 10.1016/j.joi.2009.11.010.

102. Piwowar HA, Day RS, Fridsma DB. Sharing Detailed Research Data Is Associated with Increased Citation Rate. PLoS ONE. 2007; 2: e308. DOI: 10.1371/journal.pone.0000308.

103. Rauh S, Bowers A, Rorah D, Tritz D, Pate H, Frye L, Vassar M. Evaluating the reproducibility of research in obstetrics and gynecology. European Journal of Obstetrics & Gynecology and Reproductive Biology. 2022; 269: 24–29. DOI: 10.1016/j.ejogrb.2021.12.021.

104. Rauh S, Johnson BS, Bowers A, Tritz D, Vassar BM. A review of reproducible and transparent research practices in urology publications from 2014 to2018. BMC Urology. 2022;22. DOI: 10.1186/s12894-022-01059-8.

105. Rauh S, Torgerson T, Johnson AL, Pollard J, Tritz D, Vassar M. Reproducible and transparent research practices in published neurology research. Research Integrity and Peer Review. 2020;5. DOI: 10.1186/s41073-020-0091-5.

106. Read KB, Ganshorn H, Rutley S, Scott DR. Data-sharing practices in publications funded by the Canadian Institutes of Health Research: a descriptive analysis. CMAJ Open. 2021; 9: E980–E987. DOI: 10.9778/cmajo.20200303.

107. Reidpath DD, Allotey PA. Data Sharing in Medical Research: An Empirical Investigation. Bioethics. 2001; 15: 125–134. DOI: 10.1111/1467-8519.00220.

108. Rhee S-Y, Kassaye SG, Jordan MR, Kouamou V, Katzenstein D, Shafer RW. Public availability of HIV-1 drug resistance sequence and treatment data: a systematic review. The Lancet Microbe. 2022; 3: e392–e398. DOI: 10.1016/s2666-5247(21)00250-0.

109. Riedel N, Kip M, Bobrov E. ODDPub - a Text-Mining Algorithm to Detect Data Sharing in Biomedical Publications. Data Science Journal. 2020; 19: 42. DOI: 10.5334/dsj-2020-042.

110. Rousi AM. Using current research information systems to investigate data acquisition and data sharing practices of computer scientists. Journal of Librarianship and Information Science. 2022;096100062210930. DOI: 10.1177/09610006221093049.

111. Rowhani-Farid A, Barnett AG. Badges for sharing data and code at Biostatistics: an observational study. F1000Research. 2018; 7: 90. DOI: 10.12688/f1000research.13477.2.

112. Rowhani-Farid A, Barnett AG. Has open data arrived at the British Medical Journal (BMJ)? An observational study. BMJ Open. 2016; 6: e011784. DOI: 10.1136/bmjopen-2016-011784.

113. Sarker A, Ginn R, Nikfarjam A, O’Connor K, Smith K, Jayaraman S, Upadhaya T, Gonzalez G. Utilizing social media data for pharmacovigilance: A review. Journal of Biomedical Informatics. 2015; 54: 202–212. DOI: 10.1016/j.jbi.2015.02.004.

114. Savage CJ, Vickers AJ. Empirical Study of Data Sharing by Authors Publishing in PLoS Journals. PLoS ONE. 2009; 4: e7078. DOI: 10.1371/journal.pone.0007078.

115. Schulz R, Langen G, Prill R, Cassel M, Weissgerber TL. Reporting and transparent research practices in sports medicine and orthopaedic clinical trials: a meta-research study. BMJ Open. 2022; 12: e059347. DOI: 10.1136/bmjopen-2021-059347.

116. Seibold H, Czerny S, Decke S, Dieterle R, Eder T, Fohr S, Hahn N, Hartmann R, Heindl C, Kopper P, et al. A computational reproducibility study of PLOS ONE articles featuring longitudinal data analyses. PLOS ONE. 2021; 16: e0251194. DOI: 10.1371/journal.pone.0251194.

117. Sherry CE, Pollard JZ, Tritz D, Carr BK, Pierce A, Vassar M. Assessment of transparent and reproducible research practices in the psychiatry literature. General Psychiatry. 2020; 33: e100149. DOI: 10.1136/gpsych-2019-100149.

118. Siebert M, Gaba JF, Caquelin L, Gouraud H, Dupuy A, Moher D, Naudet F. Data-sharing recommendations in biomedical journals and randomised controlled trials: an audit of journals following the ICMJE recommendations. BMJ Open. 2020; 10: e038887. DOI: 10.1136/bmjopen-2020-038887.

119. Smith CA, Nolan J, Tritz DJ, Heavener TE, Pelton J, Cook K, Vassar M. Evaluation of reproducible and transparent research practices in pulmonology. Pulmonology. 2021; 27: 134–143. DOI: 10.1016/j.pulmoe.2020.07.001.

120. Sofi-Mahmudi A, Raittio E. Transparency of COVID-19-Related Research in Dental Journals. Frontiers in Oral Health. 2022;3. DOI: 10.3389/froh.2022.871033.

121. Strcic J, Civljak A, Glozinic T, et al. Open data and data sharing in articles about COVID-19 published in preprint servers medRxiv and bioRxiv. Scientometrics. 2022; 127: 2791–2802. DOI: 10.1007/s11192-022-04346-1.

122. Sumner J, Haynes L, Nathan S, Hudson-Vitale C, McIntosh LD. Reproducibility and reporting practices in COVID-19 preprint manuscripts. 2020; DOI: 10.1101/2020.03.24.20042796.

123. Tedersoo L, Kungas R, Oras E, Koster K, Eenmaa H, Leijen A, Pedaste M, Raju M, Astapova A, Lukner H, et al. Data sharing practices and data availability upon request differ across scientific disciplines. Scientific Data. 2021;8. DOI: 10.1038/s41597-021-00981-0.

124. Thelwall M, Munafo M, Mas-Bleda A, Stuart E, Makita M, Weigert V, Keene C, Khan N, Drax K, Kousha K. Is useful research data usually shared? An investigation of genome-wide association study summary statistics. PLOS ONE. 2020; 15: e0229578. DOI: 10.1371/journal.pone.0229578.

125. Uribe SE, Sofi-Mahmudi A, Raittio E, Maldupa I, Vilne B. Dental Research Data Availability and Quality According to the FAIR Principles. Journal of Dental Research. 2022; 101: 1307–1313. DOI: 10.1177/00220345221101321.

126. Vanpaemel W, Vermorgen M, Deriemaecker L, Storms G. Are We Wasting a Good Crisis? The Availability of Psychological Research Data after the Storm. Collabra. 2015; 1. DOI: 10.1525/collabra.13.

127. Vassar M, Jellison S, Wendelbo H, Wayant C. Data sharing practices in randomized trials of addiction interventions. Addictive Behaviors. 2020; 102: 106193. DOI: 10.1016/j.addbeh.2019.106193.

128. Wallach JD, Boyack KW, Ioannidis JPA. Reproducible research practices, transparency, and open access data in the biomedical literature, 2015-2017. PLOS Biology. 2018; 16: e2006930. DOI: 10.1371/journal.pbio.2006930.

129. Walters C, Harter ZJ, Wayant C, Vo N, Warren M, Chronister J, Tritz D, Vassar M. Do oncology researchers adhere to reproducible and transparent principles? A cross-sectional survey of published oncology literature. BMJ Open. 2019; 9: e033962. DOI: 10.1136/bmjopen-2019-033962.

130. Weissgerber T, Riedel N, Kilicoglu H, Labbe C, Eckmann P, ter Riet G, Byrne J, Cabanac G, Capes-Davis A, Favier B, et al. Automated screening of COVID-19 preprints: can we help authors to improve transparency and reproducibility? Nature Medicine. 2021; 27: 6–7. DOI: 10.1038/s41591-020-01203-7.

131. Wicherts JM, Crompvoets EAV. The poor availability of syntaxes of structural equation modeling. Accountability in Research. 2017; 24: 458–468. DOI: 10.1080/08989621.2017.1396214.

132. Womack RP. Research Data in Core Journals in Biology, Chemistry, Mathematics, and Physics. PLOS ONE. 2015; 10: e0143460. DOI: 10.1371/journal.pone.0143460.

133. Wright BD, Vo N, Nolan J, Johnson AL, Braaten T, Tritz D, Vassar M. An analysis of key indicators of reproducibility in radiology. Insights into Imaging. 2020;11. DOI: 10.1186/s13244-020-00870-x.

134. Helliwell JA, Bolton WS, Burke JR, Tiernan JP, Jayne DG, Chapman SJ. Global academic response to COVID-19: Cross-sectional study. Learned Publishing. 2020; 33: 385–393. DOI: 10.1002/leap.1317.

135. Hemkens LG, Benchimol EI, Langan SM, Briel M, Kasenda B, Januel J-M, Herrett E, von Elm E. The reporting of studies using routinely collected health data was often insufficient. Journal of Clinical Epidemiology. 2016; 79: 104–111. DOI: 10.1016/j.jclinepi.2016.06.005.

136. Jiao C, Li K, Fang Z. Data sharing practices across knowledge domains: A dynamic examination of data availability statements in PLOS ONE publications. Journal of Information Science. 016555152211018;2022. DOI: 10.1177/01655515221101830.

137. McDonald L, Schultze A, Simpson A, Graham S, Wasiak R, Ramagopalan SV. A review of data sharing statements in observational studies published in the BMJ: A cross-sectional study. F1000Research. 2017; 6: 1708. DOI: 10.12688/f1000research.12673.2.

138. Ramke J, Kuper H, Limburg H, Kinloch J, Zhu W, Lansingh VC, Congdon N, Foster A, Gilbert CE. Avoidable Waste in Ophthalmic Epidemiology: A Review of Blindness Prevalence Surveys in Low and Middle Income Countries 2000-2014. Ophthalmic Epidemiology. 2017; 25: 13–20. DOI: 10.1080/09286586.2017.1328067.

139. Rustici G, Williams E, Barzine M, Brazma A, Bumgarner R, Chierici M, Furlanello C, Greger L, Jurman G, Miller M, et al. Transcriptomics data availability and reusability in the transition from microarray to next-generation sequencing. 2021; DOI: 10.1101/2020.12.31.425022.

140. Stodden V, Seiler J, Ma Z. An empirical analysis of journal policy effectiveness for computational reproducibility. Proceedings of the National Academy of Sciences. 2018; 115: 2584–2589. DOI: 10.1073/pnas.1708290115.

141. Towse JN, Ellis DA, Towse AS. Opening Pandora’s Box: Peeking inside Psychology’s data sharing practices, and seven recommendations for change. Behavior Research Methods. 2020; 53: 1455–1468. DOI: 10.3758/s13428-020-01486-1.

142. Zhao M, Yan E, Li K. Data set mentions and citations: A content analysis of full-text publications. Journal of the Association for Information Science and Technology. 2017; 69: 32–46. DOI: 10.1002/asi.23919.

143. Krawczyk M, Reuben E. (Un)Available upon Request: Field Experiment on Researchers’ Willingness to Share Supplementary Materials. Accountability in Research. 2012 May 1;19(3):175–86.

144. Minocher R, Atmaca S, Bavero C, McElreath R, Beheim B. Estimating the reproducibility of social learning research published between 1955 and 2018. Royal Society Open Science. 2021 Sep 15;8(9):210450.

145. Wicherts JM, Borsboom D, Kats J, Molenaar D. The poor availability of psychological research data for reanalysis. American Psychologist. 2006;61:726–8.

146. Vines TH, Andrew RL, Bock DG, Franklin MT, Gilbert KJ, Kane NC, et al. Mandated data archiving greatly improves access to research data. FASEB j. 2013 Apr;27(4):1304–8.

147. Vines TH, Albert AYK, Andrew RL, Débarre F, Bock DG, Franklin MT, et al. The Availability of Research Data Declines Rapidly with Article Age. Current Biology. 2014 Jan 6;24(1):94–7.

148. Roche DG, Raby GD, Norin T, Ern R, Scheuffele H, Skeeles M, et al. Paths towards greater consensus building in experimental biology. Journal of Experimental Biology. 2022 Mar 8;225(Suppl_1):jeb243559.

149. Stagge JH, Rosenberg DE, Abdallah AM, Akbar H, Attallah NA, James R. Assessing data availability and research reproducibility in hydrology and water resources. Sci Data. 2019 Feb 26;6(1):190030.

150. Tenopir C, Dalton ED, Allard S, Frame M, Pjesivac I, Birch B, et al. Changes in Data Sharing and Data Reuse Practices and Perceptions among Scientists Worldwide. PLoS One [Internet]. 2015 Aug 26;10(8).

151. Gourd E. GDPR obstructs cancer research data sharing. The Lancet Oncology. 2021 May 1;22(5):592.

152. Evans BJ, Jarvik GP. Impact of HIPAA’s minimum necessary standard on genomic data sharing. Genetics in Medicine. 2018 May 1;20(5):531–5.

153. Anagnostou P, Capocasa M, Milia N, Sanna E, Battaggia C, Luzi D, et al. When Data Sharing Gets Close to 100%: What Human Paleogenetics Can Teach the Open Science Movement. PLOS ONE. 2015 Mar 23;10(3):e0121409.

154. Thelwall M, Kousha K. Do journal data sharing mandates work? Life sciences evidence from Dryad. Aslib Journal of Information Management. 2017 Jan 1;69(1):36–45.

155. Couture JL, Blake RE, McDonald G, Ward CL. A funder-imposed data publication requirement seldom inspired data sharing. PLoS ONE. 2018 Jul 6;13(7):e0199789.

156. Rowhani-Farid A, Barnett AG. Badges for sharing data and code at Biostatistics: an observational study. F1000Research. 2018;7(101594320):90.

157. Rowhani-Farid A, Allen M, Barnett AG. What incentives increase data sharing in health and medical research? A systematic review. Research Integrity and Peer Review. 2017 May 5;2(1):4.

158. Devriendt T, Shabani M, Borry P. Data Sharing in Biomedical Sciences: A Systematic Review of Incentives. Biopreservation and Biobanking. 2021 Jun;19(3):219–27.

159. Institute of Medicine (IOM). 2015. Sharing clinical trial data: Maximizing benefits, minimizing risk. Washington, DC: The National Academies Press.

160. Cristea IA, Naudet F, Caquelin L. Meta-research studies should improve and evaluate their own data sharing practices. Journal of Clinical Epidemiology. 2022 Sep 1;149:183–9.

161. Migliavaca CB, Stein C, Colpani V, Barker TH, Ziegelmann PK, Munn Z, et al. Meta-analysis of prevalence: I2 statistic and how to deal with heterogeneity. Research Synthesis Methods. 2022;13(3):363–7.

162. Jia P, Lin L, Kwong JSW, Xu C. Many metaanalyses of rare events in the Cochrane Database of Systematic Reviews were underpowered. Journal of Clinical Epidemiology. 2021 Mar 1;131:113–22.

163. Deeks JJ, Higgins JPT, Altman DG: Chapter 10: Analysing data and undertaking meta-analyses. In: Higgins JPT, Thomas J, Chandler J, et al. (editors). Cochrane Handbook for Systematic Reviews of Interventions version 6.3. Cochrane; 2022.

164. Warton DI, Hui FKC. The arcsine is asinine: the analysis of proportions in ecology. Ecology. 2011;92(1):3–10.

165. Xu C, Li L, Lin L, Chu H, Thabane L, Zou K, et al. Exclusion of studies with no events in both arms in meta-analysis impacted the conclusions. Journal of Clinical Epidemiology. 2020 Jul 1;123:91–9.

166. Smith-Spangler CM. Transparency and Reproducible Research in Modeling: Why We Need It and How to Get There. Med Decis Making. 2012 Sep 1;32(5):663–6.

167. Dewey M, Bosserdt M, Dodd JD, Thun S, Kressel HY. Clinical Imaging Research: Higher Evidence, Global Collaboration, Improved Reporting, and Data Sharing Are the Grand Challenges. Radiology. 2019 Jun;291(3):547–52.

168. Spidlen J, Brinkman RR. Use FlowRepository to share your clinical data upon study publication: USING FlowRepository FOR CLINICAL PUBLICATIONS. Cytometry. 2018 Jan;94(1):196–8.

169. Bisson J, Simmler C, Chen SN, Friesen JB, Lankin DC, McAlpine JB, et al. Dissemination of original NMR data enhances reproducibility and integrity in chemical research. Nat Prod Rep. 2016;33(9):1028–33.

170. Helliwell JR, Minor W, Weiss MS, Garman EF, Read RJ, Newman J, et al. Findable Accessible Interoperable Re-usable (FAIR) diffraction data are coming to protein crystallography. Acta Cryst F. 2019 May 1;75(5):321–3.

171. Kroon-Batenburg Lmj, Helliwell JR, McMahon B, Terwilliger TC. Raw diffraction data preservation and reuse: overview, update on practicalities and metadata requirements. IUCrJ. 2017 Jan 1;4(1):87–99.

172. Antes AL, Walsh HA, Strait M, Hudson-Vitale CR, DuBois JM. Examining Data Repository Guidelines for Qualitative Data Sharing. Journal of Empirical Research on Human Research Ethics. 2018 Feb 1;13(1):61–73.

173. Campbell HA, Micheli-Campbell MA, Udyawer V. Early Career Researchers Embrace Data Sharing. Trends in Ecology & Evolution. 2019 Feb 1;34(2):95–8.

