## Supplementary material for "Rates and predictors of data and code sharing in the medical and health sciences: A systematic review with meta-analysis of individual participant data"

### SUPPLEMENTARY FIGURES AND TABLES

#### Supplementary Figure 1. Summary of the results of the risk of bias assessments.

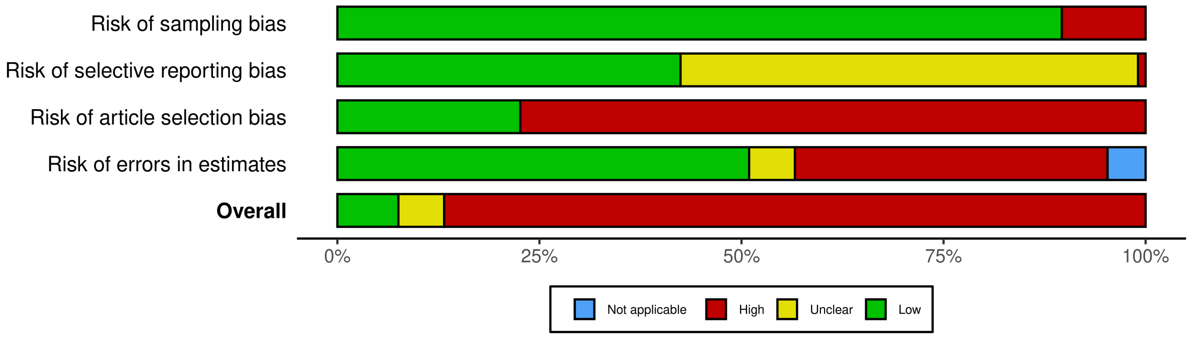

**Supplementary Figure 2. Risk of bias traffic light plot.**

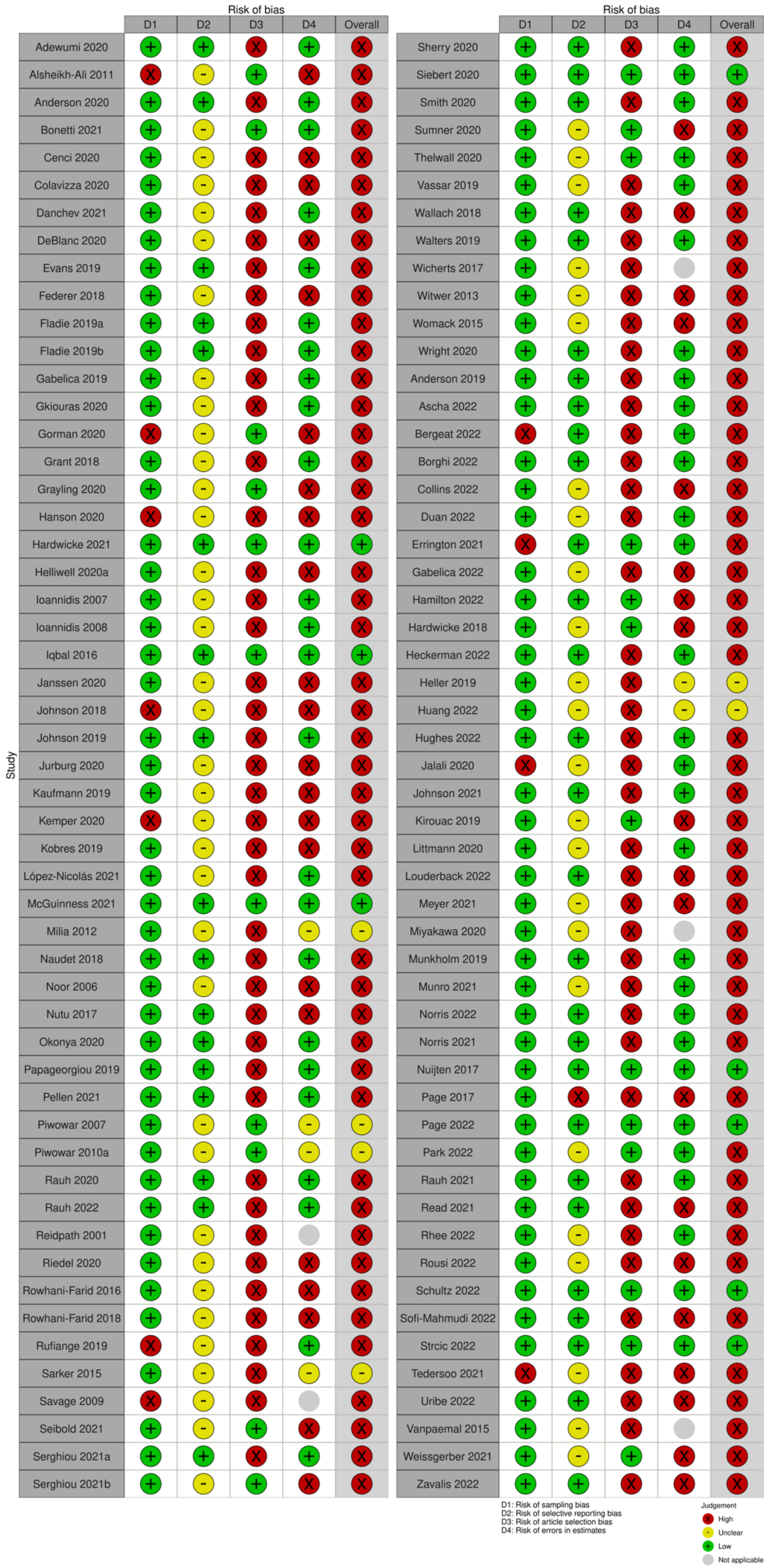

**Supplementary Figure 3. Proportion of primary articles assessed by eligible meta-research studies flagged as potential duplicates.**

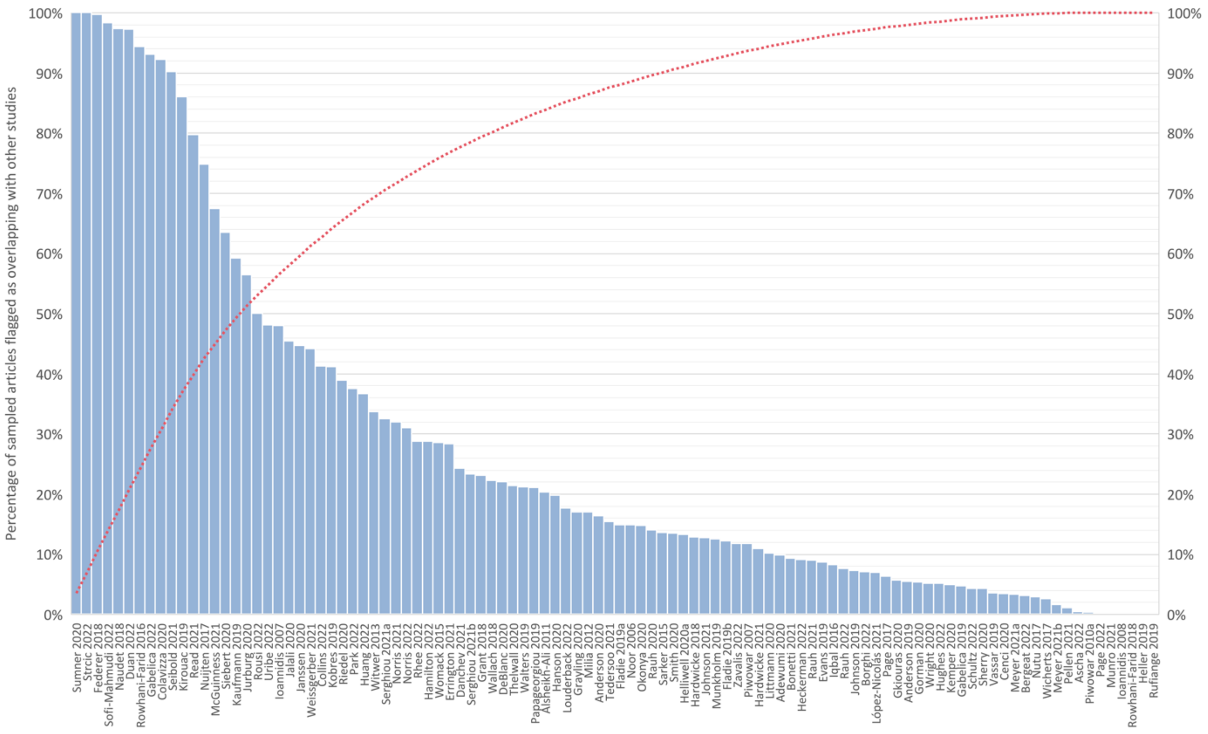

**Supplementary Figure 4. Declared private data sharing rates since 2016.**

| Study | Source | Coding | ROB | Events | Total | Proportion | 95%–CI | Weight (random) |
|---|---|---|---|---|---|---|---|---|
| Hardwicke 2021 | IPD | Manual | Low | 0 | 49 | 0.00 | [0.00; 0.07] | 3.2% |
| Rauh 2022 | IPD | Manual | High | 0 | 113 | 0.00 | [0.00; 0.03] | 4.3% |
| Wright 2020 | IPD | Manual | High | 0 | 108 | 0.00 | [0.00; 0.03] | 4.2% |
| Nutu 2017 | IPD | Manual | High | 1 | 203 | 0.00 | [0.00; 0.03] | 4.8% |
| Ascha 2022 | IPD | Manual | High | 1 | 159 | 0.01 | [0.00; 0.03] | 4.6% |
| Hughes 2022 | IPD | Manual | High | 1 | 122 | 0.01 | [0.00; 0.04] | 4.4% |
| Anderson 2019 | IPD | Manual | High | 1 | 83 | 0.01 | [0.00; 0.07] | 3.9% |
| Okonya 2020 | IPD | Manual | High | 1 | 71 | 0.01 | [0.00; 0.08] | 3.7% |
| Wallach 2018 | IPD | Manual | High | 1 | 52 | 0.02 | [0.00; 0.10] | 3.3% |
| Johnson 2019 | IPD | Manual | High | 2 | 103 | 0.02 | [0.01; 0.07] | 4.2% |
| Adewumi 2020 | IPD | Manual | High | 3 | 154 | 0.02 | [0.01; 0.06] | 4.6% |
| Smith 2020 | IPD | Manual | High | 3 | 138 | 0.02 | [0.01; 0.06] | 4.5% |
| Johnson 2021 | IPD | Manual | High | 3 | 128 | 0.02 | [0.01; 0.07] | 4.4% |
| Anderson 2020 | IPD | Manual | High | 4 | 150 | 0.03 | [0.01; 0.07] | 4.6% |
| Rauh 2020 | IPD | Manual | High | 5 | 168 | 0.03 | [0.01; 0.07] | 4.7% |
| Evans 2019 | IPD | Manual | High | 4 | 133 | 0.03 | [0.01; 0.07] | 4.5% |
| Fladie 2019a | IPD | Manual | High | 5 | 121 | 0.04 | [0.02; 0.09] | 4.4% |
| Sherry 2020 | IPD | Manual | High | 6 | 140 | 0.04 | [0.02; 0.09] | 4.5% |
| Rauh 2021 | IPD | Manual | High | 6 | 129 | 0.05 | [0.02; 0.10] | 4.4% |
| Serghiou 2021a | IPD | Manual | High | 11 | 207 | 0.05 | [0.03; 0.09] | 4.9% |
| Fladie 2019b | IPD | Manual | High | 6 | 106 | 0.06 | [0.03; 0.12] | 4.2% |
| Walters 2019 | IPD | Manual | High | 10 | 116 | 0.09 | [0.05; 0.15] | 4.3% |
| Hamilton 2022 | IPD | Manual | High | 38 | 305 | 0.12 | [0.09; 0.17] | 5.1% |
| **Random effects (GLMM method)** | | | | | **3058** | **0.02** | **[0.02; 0.04]** | **--** |
| **Random effects (HKSJ method)** | | | | | | **0.02** | **[0.01; 0.04]** | **100.0%** |
| **Prediction interval** | | | | | | | **[0.00; 0.10]** | |

Heterogeneity: $I^2 = 80\%$, $\tau^2 < 0.01$, $p < 0.01$

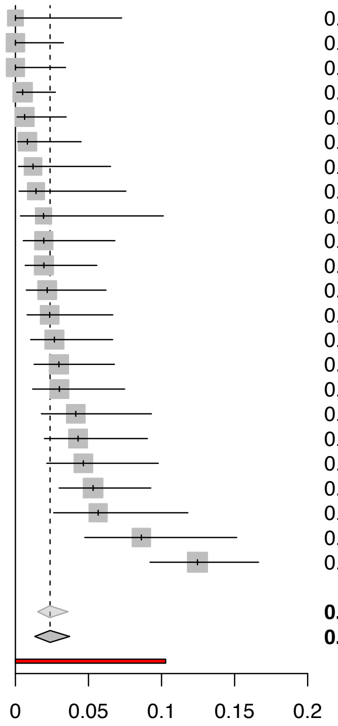

#### Supplementary Figure 5. Declared private code sharing rates since 2016.

| Study | Source | Coding | ROB | Events | Total | Proportion | 95%–CI | Weight (random) |
|---|---|---|---|---|---|---|---|---|
| Adewumi 2020 | IPD | Manual | High | 0 | 154 | 0.00 | [0.00; 0.02] | 5.4% |
| Evans 2019 | IPD | Manual | High | 0 | 133 | 0.00 | [0.00; 0.03] | 4.8% |
| Fladie 2019a | IPD | Manual | High | 0 | 121 | 0.00 | [0.00; 0.03] | 4.4% |
| Fladie 2019b | IPD | Manual | High | 0 | 106 | 0.00 | [0.00; 0.03] | 3.9% |
| Hardwicke 2021 | IPD | Manual | Low | 0 | 49 | 0.00 | [0.00; 0.07] | 2.0% |
| Johnson 2019 | IPD | Manual | High | 0 | 103 | 0.00 | [0.00; 0.04] | 3.8% |
| Okonya 2020 | IPD | Manual | High | 0 | 72 | 0.00 | [0.00; 0.05] | 2.8% |
| Rauh 2020 | IPD | Manual | High | 0 | 168 | 0.00 | [0.00; 0.02] | 5.8% |
| Rauh 2022 | IPD | Manual | High | 0 | 113 | 0.00 | [0.00; 0.03] | 4.2% |
| Smith 2020 | IPD | Manual | High | 0 | 138 | 0.00 | [0.00; 0.03] | 4.9% |
| Wallach 2018 | IPD | Manual | High | 0 | 52 | 0.00 | [0.00; 0.07] | 2.1% |
| Walters 2019 | IPD | Manual | High | 0 | 116 | 0.00 | [0.00; 0.03] | 4.3% |
| Wright 2020 | IPD | Manual | High | 0 | 108 | 0.00 | [0.00; 0.03] | 4.0% |
| Anderson 2019 | IPD | Manual | High | 0 | 83 | 0.00 | [0.00; 0.04] | 3.2% |
| Ascha 2022 | IPD | Manual | High | 0 | 159 | 0.00 | [0.00; 0.02] | 5.5% |
| Hughes 2022 | IPD | Manual | High | 0 | 122 | 0.00 | [0.00; 0.03] | 4.4% |
| Johnson 2021 | IPD | Manual | High | 0 | 128 | 0.00 | [0.00; 0.03] | 4.6% |
| Rauh 2021 | IPD | Manual | High | 0 | 129 | 0.00 | [0.00; 0.03] | 4.7% |
| Serghiou 2021a | IPD | Manual | High | 1 | 207 | 0.00 | [0.00; 0.03] | 6.8% |
| Anderson 2020 | IPD | Manual | High | 1 | 150 | 0.01 | [0.00; 0.04] | 5.3% |
| Sherry 2020 | IPD | Manual | High | 1 | 140 | 0.01 | [0.00; 0.04] | 5.0% |
| Hamilton 2022 | IPD | Manual | High | 2 | 274 | 0.01 | [0.00; 0.03] | 8.3% |
| **Random effects (GLMM method)** | | | | | **2825** | **0.00** | **[0.00; 0.00]** | **--** |
| **Random effects (HKSJ method)** | | | | | | **0.00** | **[0.00; 0.00]** | **100.0%** |
| **Prediction interval** | | | | | | | **[0.00; 0.00]** | |

Heterogeneity: $I^2 = 0\%$, $\tau^2 < 0.01$, $p = 0.84$

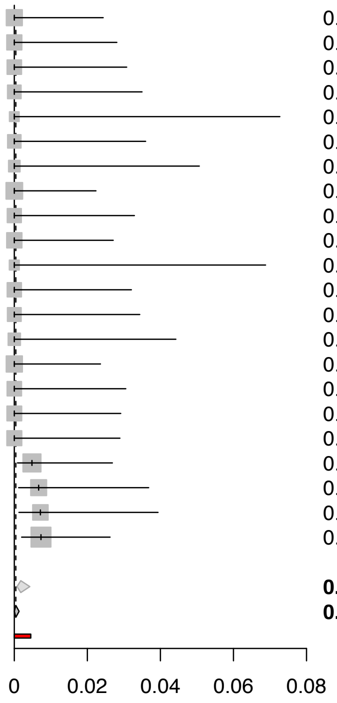

**Supplementary Figure 6. Success rates of private requests for data from published medical research by declaration type.**

| Study | Journal policy | Data type | Source | Events | Total | Proportion | 95%−CI |
|---|---|---|---|---|---|---|---|
| **Available on request** | | | | | | | |
| Milia 2012 | Mixed | Sequence data | IPD | 0 | 1 | 0.00 | [0.00; 0.79] |
| Bergeat 2022 | No policy | Trial data | IPD | 0 | 5 | 0.00 | [0.00; 0.43] |
| Gabelica 2019 | Mixed | Trial data | Paper | 1 | 24 | 0.04 | [0.01; 0.20] |
| Gabelica 2022 | Not reported | No restrictions | Author | 102 | 1540 | 0.07 | [0.05; 0.08] |
| Rowhani−Farid 2016 | Share on request | No restrictions | IPD | 4 | 32 | 0.12 | [0.05; 0.28] |
| Naudet 2018 | Share on request | Trial data | IPD | 10 | 23 | 0.43 | [0.26; 0.63] |
| Tedersoo 2021 | Mixed | No restrictions | IPD | 2 | 2 | 1.00 | [0.34; 1.00] |
| Stodden 2018 | No restrictions | Share on request | NA | NA | NA | NA | [NA; NA] |
| Heterogeneity: $I^2 = 83\%$, $\tau^2 = 0.22$, $p < 0.01$ | | | | | | | |
| **No availability statement** | | | | | | | |
| Naudet 2018 | Share on request | Trial data | IPD | 0 | 1 | 0.00 | [0.00; 0.79] |
| Bergeat 2022 | No policy | Trial data | IPD | 4 | 119 | 0.03 | [0.01; 0.08] |
| Reidpath 2001 | No policy | No restrictions | Paper | 1 | 29 | 0.03 | [0.01; 0.17] |
| Gabelica 2019 | Mixed | Trial data | Paper | 23 | 595 | 0.04 | [0.03; 0.06] |
| Hardwicke 2018 | Not reported | No restrictions | IPD | 7 | 73 | 0.10 | [0.05; 0.18] |
| Savage 2009 | Share on request | No restrictions | Paper | 1 | 10 | 0.10 | [0.02; 0.40] |
| Tedersoo 2021 | Mixed | No restrictions | IPD | 7 | 23 | 0.30 | [0.16; 0.51] |
| Heterogeneity: $I^2 = 65\%$, $\tau^2 = 0.02$, $p < 0.01$ | | | | | | | |
| **Stated as not available** | | | | | | | |
| Naudet 2018 | Share on request | Trial data | IPD | 0 | 3 | 0.00 | [0.00; 0.56] |
| Bergeat 2022 | No policy | Trial data | IPD | 0 | 6 | 0.00 | [0.00; 0.39] |
| Heterogeneity: $I^2 = 0\%$, $\tau^2 = 0$, $p = 1.00$ | | | | | | | |

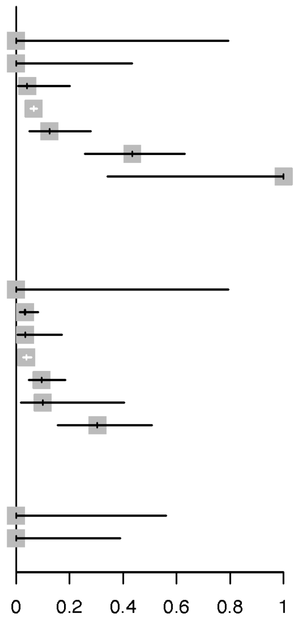

**Supplementary Figure 7. Declared and actual public code sharing rates by journal code sharing policy.**

| Study | Code type | Source | Events | Total | Proportion | 95%-CI |
|---|---|---|---|---|---|---|
| **Reported code sharing: No sharing policy** | | | | | | |
| Hamilton 2022 | No restrictions | IPD | 4 | 179 | 0.02 | [0.01; 0.06] |
| Grayling 2020 | Methods code | IPD | 8 | 48 | 0.17 | [0.09; 0.30] |
| Littmann 2020 | No restrictions | IPD | 9 | 53 | 0.17 | [0.09; 0.29] |
| Heterogeneity: $I^2 = 90\%$, $\tau^2 = 0.02$, $p < 0.01$ | | | | | | |
| **Reported code sharing: 'Encourage' policy** | | | | | | |
| Hamilton 2022 | No restrictions | IPD | 3 | 81 | 0.04 | [0.01; 0.10] |
| Grayling 2020 | Methods code | IPD | 43 | 106 | 0.41 | [0.32; 0.50] |
| Littmann 2020 | No restrictions | IPD | 16 | 30 | 0.53 | [0.36; 0.70] |
| Heterogeneity: $I^2 = 97\%$, $\tau^2 = 0.10$, $p < 0.01$ | | | | | | |
| **Reported code sharing: Mandatory sharing policy** | | | | | | |
| Hamilton 2022 | No restrictions | IPD | 1 | 8 | 0.12 | [0.02; 0.47] |
| Grayling 2020 | Methods code | IPD | 28 | 91 | 0.31 | [0.22; 0.41] |
| Littmann 2020 | No restrictions | IPD | 26 | 47 | 0.55 | [0.41; 0.69] |
| Heterogeneity: $I^2 = 81\%$, $\tau^2 = 0.04$, $p < 0.01$ | | | | | | |
| **Actual code sharing: No sharing policy** | | | | | | |
| Grayling 2020 | Methods code | IPD | 8 | 48 | 0.17 | [0.09; 0.30] |
| **Actual code sharing: 'Encourage' policy** | | | | | | |
| Grayling 2020 | Methods code | IPD | 29 | 106 | 0.27 | [0.20; 0.37] |
| **Actual code sharing: Mandatory sharing policy** | | | | | | |
| Grayling 2020 | Methods code | IPD | 16 | 91 | 0.18 | [0.11; 0.27] |
| Kirouac 2019 | Model data | IPD | 18 | 22 | 0.82 | [0.61; 0.93] |
| Heterogeneity: $I^2 = 97\%$, $\tau^2 = 0.23$, $p < 0.01$ | | | | | | |

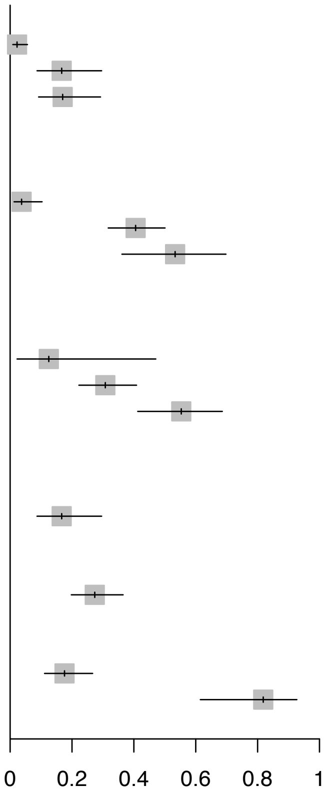

### Supplementary Figure 8. Association between data sharing and code sharing (actual availability).

| | | | | Data sharers | | Data witholders | | | | |
|---|---|---|---|---|---|---|---|---|---|---|
| Study | Source | Coding | ROB | Events | Total | Events | Total | RR | 95%−CI | Weight (random) |
| Anderson 2020 | IPD | Manual | High | 0 | 8 | 2 | 224 | 0.33 | [ 0.00; 12970.39] | 2.0% |
| Smith 2020 | IPD | Manual | High | 0 | 4 | 1 | 241 | 0.50 | [ 0.00; 2348130.65] | 1.0% |
| Rauh 2020 | IPD | Manual | High | 0 | 2 | 1 | 269 | 0.50 | [ 0.00; 4033891717.70] | 0.4% |
| Riedel 2020 | IPD | Manual | High | 6 | 72 | 4 | 980 | 20.42 | [ 5.89; 70.72] | 31.3% |
| Adewumi 2020 | IPD | Manual | High | 2 | 12 | 0 | 231 | 41.50 | [ 3.91; 440.50] | 20.4% |
| Fladie 2019b | IPD | Manual | High | 3 | 11 | 1 | 161 | 43.91 | [ 4.97; 388.12] | 22.0% |
| Louderback 2022 | IPD | Manual | High | 7 | 16 | 0 | 483 | 219.31 | [27.79; 1730.69] | 23.0% |
| Evans 2019 | IPD | Manual | High | 0 | 1 | 0 | 194 | | | 0.0% |
| Fladie 2019a | IPD | Manual | High | 0 | 2 | 0 | 180 | | | 0.0% |
| Okonya 2020 | IPD | Manual | High | 0 | 1 | 0 | 121 | | | 0.0% |
| Sherry 2020 | IPD | Manual | High | 0 | 1 | 0 | 210 | | | 0.0% |
| Walters 2019 | IPD | Manual | High | 0 | 3 | 0 | 191 | | | 0.0% |
| Wright 2020 | IPD | Manual | High | 0 | 1 | 0 | 193 | | | 0.0% |
| Anderson 2019 | IPD | Manual | High | 0 | 3 | 0 | 124 | | | 0.0% |
| Ascha 2022 | IPD | Manual | High | 0 | 2 | 0 | 201 | | | 0.0% |
| Hughes 2022 | IPD | Manual | High | 0 | 6 | 0 | 190 | | | 0.0% |
| Johnson 2021 | IPD | Manual | High | 0 | 2 | 0 | 210 | | | 0.0% |
| **Random effects (BGLMM method)** | | | | | **147** | | **4403** | **40.42** | **[15.45; 120.39]** | **--** |
| **Random effects (HKSJ method)** | | | | | | | | **42.05** | **[12.15; 145.52]** | **100.0%** |
| **Prediction interval** | | | | | | | | | **[ 0.94; 1879.62]** | |

Heterogeneity: $I^2 = 0\%$, $\tau^2 = 1.57$, $p = 0.55$

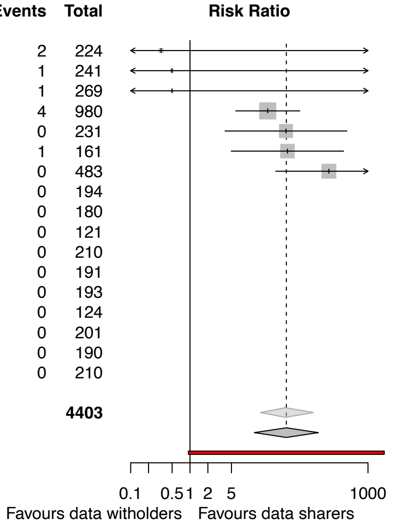

**Supplementary Figure 9. Declared public data sharing rates since 2016 by data type.**

| Study | Source | Coding | ROB | Events | Total | Proportion | 95%−CI | Weight (random) |
|---|---|---|---|---|---|---|---|---|
| **No restrictions** | | | | | | | | |
| Hardwicke 2021 | IPD | Manual | Low | 0 | 49 | 0.00 | [0.00; 0.07] | 3.3% |
| Sofi−Mahmudi 2022 | IPD | Auto | High | 0 | 11 | 0.00 | [0.00; 0.26] | 2.0% |
| Johnson 2019 | IPD | Manual | High | 1 | 103 | 0.01 | [0.00; 0.05] | 3.7% |
| Nutu 2017 | IPD | Manual | High | 3 | 203 | 0.01 | [0.01; 0.04] | 3.9% |
| Ascha 2022 | IPD | Manual | High | 4 | 159 | 0.03 | [0.01; 0.06] | 3.9% |
| Uribe 2022 | IPD | Both | High | 197 | 7456 | 0.03 | [0.02; 0.03] | 4.1% |
| Sherry 2020 | IPD | Manual | High | 4 | 140 | 0.03 | [0.01; 0.07] | 3.8% |
| Evans 2019 | IPD | Manual | High | 5 | 133 | 0.04 | [0.02; 0.08] | 3.8% |
| Rauh 2022 | IPD | Manual | High | 5 | 113 | 0.04 | [0.02; 0.10] | 3.7% |
| Fladie 2019a | IPD | Manual | High | 6 | 121 | 0.05 | [0.02; 0.10] | 3.8% |
| Rauh 2020 | IPD | Manual | High | 9 | 168 | 0.05 | [0.03; 0.10] | 3.9% |
| Walters 2019 | IPD | Manual | High | 8 | 116 | 0.07 | [0.04; 0.13] | 3.8% |
| Serghiou 2021a | IPD | Manual | High | 15 | 206 | 0.07 | [0.04; 0.12] | 3.9% |
| Wright 2020 | IPD | Manual | High | 8 | 108 | 0.07 | [0.04; 0.14] | 3.7% |
| Serghiou 2021b | IPD | Auto | High | 53526 | 689457 | 0.08 | [0.08; 0.08] | 4.1% |
| Johnson 2021 | IPD | Manual | High | 10 | 128 | 0.08 | [0.04; 0.14] | 3.8% |
| Rauh 2021 | IPD | Manual | High | 11 | 129 | 0.09 | [0.05; 0.15] | 3.8% |
| Hughes 2022 | IPD | Manual | High | 11 | 122 | 0.09 | [0.05; 0.15] | 3.8% |
| Adewumi 2020 | IPD | Manual | High | 14 | 154 | 0.09 | [0.05; 0.15] | 3.8% |
| Anderson 2019 | IPD | Manual | High | 10 | 83 | 0.12 | [0.07; 0.21] | 3.6% |
| Smith 2020 | IPD | Manual | High | 21 | 138 | 0.15 | [0.10; 0.22] | 3.8% |
| Wallach 2018 | IPD | Manual | High | 8 | 52 | 0.15 | [0.08; 0.28] | 3.4% |
| Okonya 2020 | IPD | Manual | High | 13 | 71 | 0.18 | [0.11; 0.29] | 3.5% |
| Hamilton 2022 | IPD | Manual | High | 59 | 305 | 0.19 | [0.15; 0.24] | 4.0% |
| Anderson 2020 | IPD | Manual | High | 30 | 150 | 0.20 | [0.14; 0.27] | 3.8% |
| Fladie 2019b | IPD | Manual | High | 25 | 106 | 0.24 | [0.17; 0.33] | 3.7% |
| Kobres 2019 | Paper | Manual | High | 29 | 73 | 0.40 | [0.29; 0.51] | 3.6% |
| **Random effects (GLMM method)** | | | | | **700054** | **0.07** | **[0.05; 0.10]** | **−−** |
| **Random effects (HKSJ method)** | | | | | | **0.08** | **[0.05; 0.11]** | **100.0%** |
| **Prediction interval** | | | | | | | **[0.00; 0.30]** | |
| Heterogeneity: $I^2 = 96\%$, $\tau^2 = 0.02$, $p < 0.01$ | | | | | | | | |
| **Review data** | | | | | | | | |
| Page 2022 | IPD | Manual | Low | 33 | 297 | 0.11 | [0.08; 0.15] | 54.2% |
| López−Nicolás 2021 | IPD | Manual | High | 18 | 53 | 0.34 | [0.23; 0.47] | 45.8% |
| **Random effects (GLMM method)** | | | | | **350** | **0.19** | **[0.08; 0.39]** | **−−** |
| **Random effects (HKSJ method)** | | | | | | **0.21** | **[0.00; 1.00]** | **100.0%** |
| Heterogeneity: $I^2 = 93\%$, $\tau^2 = 0.03$, $p < 0.01$ | | | | | | | | |
| **Trial data** | | | | | | | | |
| Duan 2022 | IPD | Manual | High | 0 | 2 | 0.00 | [0.00; 0.66] | 3.6% |
| Papageorgiou 2019 | IPD | Manual | High | 15 | 299 | 0.05 | [0.03; 0.08] | 24.9% |
| Norris 2022 | IPD | Manual | High | 5 | 99 | 0.05 | [0.02; 0.11] | 23.1% |
| Norris 2021 | IPD | Manual | High | 6 | 100 | 0.06 | [0.03; 0.12] | 23.1% |
| Borghi 2022 | IPD | Manual | High | 37 | 528 | 0.07 | [0.05; 0.10] | 25.3% |
| **Random effects (GLMM method)** | | | | | **1028** | **0.06** | **[0.05; 0.08]** | **−−** |
| **Random effects (HKSJ method)** | | | | | | **0.06** | **[0.04; 0.08]** | **100.0%** |
| **Prediction interval** | | | | | | | **[0.01; 0.14]** | |
| Heterogeneity: $I^2 = 0\%$, $\tau^2 = < 0.01$, $p = 0.72$ | | | | | | | | |
| **Model data** | | | | | | | | |
| Zavalis 2022 | IPD | Auto | High | 332 | 1336 | 0.25 | [0.23; 0.27] | 100.0% |

Test for subgroup differences (random effects): $\chi^2_3 = 229.65$, df = 3 ($p < 0.01$)

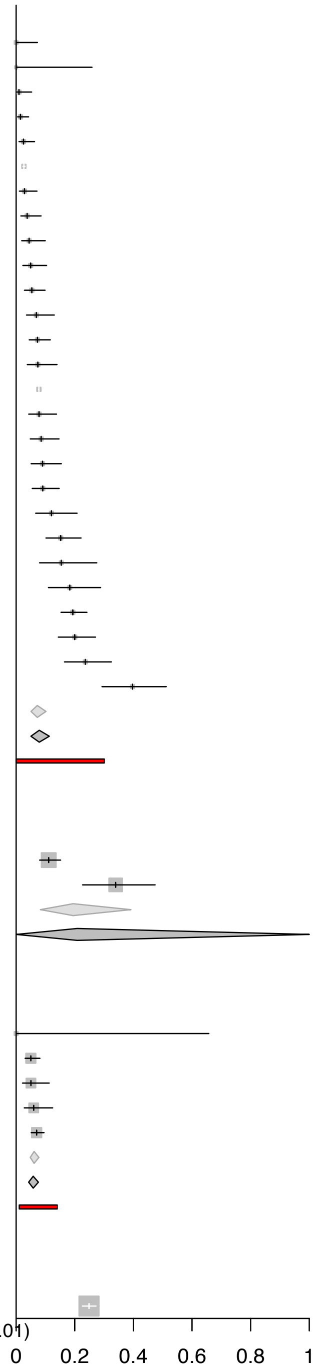

**Supplementary Figure 10. Actual public data sharing rates since 2016 by data type.**

| Study | Source | Coding | ROB | Events | Total | Proportion | 95%−CI | Weight (random) |
|---|---|---|---|---|---|---|---|---|
| **No restrictions** | | | | | | | | |
| Hardwicke 2021 | IPD | Manual | Low | 0 | 49 | 0.00 | [0.00; 0.07] | 3.8% |
| Johnson 2019 | IPD | Manual | High | 0 | 103 | 0.00 | [0.00; 0.04] | 4.0% |
| Rauh 2022 | IPD | Manual | High | 0 | 113 | 0.00 | [0.00; 0.03] | 4.0% |
| Sherry 2020 | IPD | Manual | High | 0 | 140 | 0.00 | [0.00; 0.03] | 4.1% |
| Wright 2020 | IPD | Manual | High | 0 | 108 | 0.00 | [0.00; 0.03] | 4.0% |
| Rauh 2021 | IPD | Manual | High | 0 | 129 | 0.00 | [0.00; 0.03] | 4.0% |
| Sofi−Mahmudi 2022 | IPD | Auto | High | 0 | 11 | 0.00 | [0.00; 0.26] | 3.0% |
| Rauh 2020 | IPD | Manual | High | 1 | 168 | 0.01 | [0.00; 0.03] | 4.1% |
| Evans 2019 | IPD | Manual | High | 1 | 133 | 0.01 | [0.00; 0.04] | 4.0% |
| Johnson 2021 | IPD | Manual | High | 1 | 128 | 0.01 | [0.00; 0.04] | 4.0% |
| Fladie 2019a | IPD | Manual | High | 1 | 121 | 0.01 | [0.00; 0.05] | 4.0% |
| Ascha 2022 | IPD | Manual | High | 2 | 159 | 0.01 | [0.00; 0.04] | 4.1% |
| Okonya 2020 | IPD | Manual | High | 1 | 72 | 0.01 | [0.00; 0.07] | 3.9% |
| Smith 2020 | IPD | Manual | High | 2 | 138 | 0.01 | [0.00; 0.05] | 4.0% |
| Uribe 2022 | IPD | Both | High | 112 | 7506 | 0.01 | [0.01; 0.02] | 4.2% |
| Walters 2019 | IPD | Manual | High | 2 | 116 | 0.02 | [0.00; 0.06] | 4.0% |
| Hughes 2022 | IPD | Manual | High | 3 | 122 | 0.02 | [0.01; 0.07] | 4.0% |
| Louderback 2022 | IPD | Manual | High | 16 | 499 | 0.03 | [0.02; 0.05] | 4.1% |
| Anderson 2019 | IPD | Manual | High | 3 | 83 | 0.04 | [0.01; 0.10] | 4.0% |
| Anderson 2020 | IPD | Manual | High | 6 | 150 | 0.04 | [0.02; 0.08] | 4.1% |
| Serghiou 2021a | IPD | Manual | High | 10 | 207 | 0.05 | [0.03; 0.09] | 4.1% |
| Adewumi 2020 | IPD | Manual | High | 8 | 154 | 0.05 | [0.03; 0.10] | 4.1% |
| Fladie 2019b | IPD | Manual | High | 7 | 106 | 0.07 | [0.03; 0.13] | 4.0% |
| Riedel 2020 | IPD | Manual | High | 72 | 1052 | 0.07 | [0.05; 0.09] | 4.2% |
| Hamilton 2022 | IPD | Manual | High | 49 | 306 | 0.16 | [0.12; 0.21] | 4.1% |
| **Random effects (GLMM method)** | | | | | **11873** | **0.02** | **[0.01; 0.03]** | **--** |
| **Random effects (HKSJ method)** | | | | | | **0.02** | **[0.01; 0.03]** | **100.0%** |
| **Prediction interval** | | | | | | | **[0.00; 0.11]** | |
| Heterogeneity: $I^2 = 90\%$, $\tau^2 = < 0.01$, $p < 0.01$ | | | | | | | | |
| **Review data** | | | | | | | | |
| Page 2022 | IPD | Manual | Low | 12 | 297 | 0.04 | [0.02; 0.07] | 51.1% |
| Bonetti 2021 | IPD | Manual | High | 8 | 75 | 0.11 | [0.06; 0.20] | 48.9% |
| **Random effects (GLMM method)** | | | | | **372** | **0.06** | **[0.03; 0.12]** | **--** |
| **Random effects (HKSJ method)** | | | | | | **0.06** | **[0.00; 0.77]** | **100.0%** |
| Heterogeneity: $I^2 = 75\%$, $\tau^2 = < 0.01$, $p = 0.04$ | | | | | | | | |
| **Trial data** | | | | | | | | |
| Duan 2022 | IPD | Manual | High | 0 | 36 | 0.00 | [0.00; 0.10] | 31.7% |
| Norris 2022 | IPD | Manual | High | 1 | 99 | 0.01 | [0.00; 0.06] | 34.1% |
| Norris 2021 | IPD | Manual | High | 2 | 100 | 0.02 | [0.01; 0.07] | 34.2% |
| **Random effects (GLMM method)** | | | | | **235** | **0.01** | **[0.00; 0.04]** | **--** |
| **Random effects (HKSJ method)** | | | | | | **0.01** | **[0.00; 0.06]** | **100.0%** |
| Heterogeneity: $I^2 = 6\%$, $\tau^2 = < 0.01$, $p = 0.34$ | | | | | | | | |
| **Sequence data** | | | | | | | | |
| Huang 2022 | IPD | Unclear | Unclear | 40 | 93 | 0.43 | [0.33; 0.53] | 34.0% |
| Rhee 2022 | IPD | Manual | High | 158 | 321 | 0.49 | [0.44; 0.55] | 35.1% |
| Meyer 2021a | IPD | Manual | High | 24 | 30 | 0.80 | [0.63; 0.90] | 30.9% |
| **Random effects (GLMM method)** | | | | | **444** | **0.56** | **[0.38; 0.73]** | **--** |
| **Random effects (HKSJ method)** | | | | | | **0.57** | **[0.12; 0.96]** | **100.0%** |
| Heterogeneity: $I^2 = 86\%$, $\tau^2 = 0.04$, $p < 0.01$ | | | | | | | | |

Test for subgroup differences (random effects): $\chi^2_3 = 41.43$, df = 3 ($p < 0.01$)

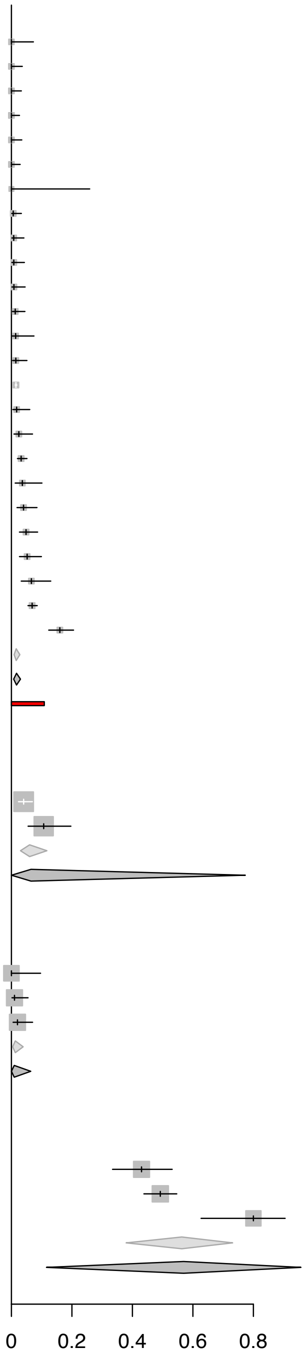

**Supplementary Figure 11. Data and code sharing rates among studies investigating COVID-19.**

| Study | Data type | Source | Coding | Events | Total | Proportion | 95%−CI | Weight (random) |
|---|---|---|---|---|---|---|---|---|
| **Reported data sharing rates** | | | | | | | | |
| Sofi−Mahmudi 2022 | No restrictions | IPD | Auto | 0 | 11 | 0.00 | [0.00; 0.26] | 23.0% |
| Weissgerber 2021 | No restrictions | IPD | Auto | 587 | 4243 | 0.14 | [0.13; 0.15] | 38.5% |
| Collins 2022 | No restrictions | IPD | Auto | 655 | 3550 | 0.18 | [0.17; 0.20] | 38.5% |
| **Random effects (BGLMM method)** | | | | | **7804** | **0.16** | **[0.13; 0.19]** | **−−** |
| **Random effects (HKSJ method)** | | | | | | **0.09** | **[0.00; 0.57]** | **100.0%** |
| Heterogeneity: $I^2 = 95\%$, $\tau^2 = 0.04$, $p < 0.01$ | | | | | | | | |
| **Actual data sharing rates** | | | | | | | | |
| Sofi−Mahmudi 2022 | No restrictions | IPD | Auto | 0 | 11 | 0.00 | [0.00; 0.26] | 24.7% |
| Strcic 2022 | No restrictions | IPD | Manual | 123 | 888 | 0.14 | [0.12; 0.16] | 41.1% |
| Gkiouras 2020 | No restrictions | IPD | Manual | 11 | 35 | 0.31 | [0.19; 0.48] | 34.2% |
| **Random effects (BGLMM method)** | | | | | **934** | **0.16** | **[0.09; 0.29]** | **−−** |
| **Random effects (HKSJ method)** | | | | | | **0.11** | **[0.00; 0.76]** | **100.0%** |
| Heterogeneity: $I^2 = 84\%$, $\tau^2 = 0.07$, $p < 0.01$ | | | | | | | | |
| **Reported code sharing rates** | | | | | | | | |
| Sofi−Mahmudi 2022 | No restrictions | IPD | Auto | 0 | 11 | 0.00 | [0.00; 0.26] | 22.9% |
| Weissgerber 2021 | No restrictions | IPD | Auto | 757 | 5667 | 0.13 | [0.12; 0.14] | 38.5% |
| Collins 2022 | No restrictions | IPD | Auto | 654 | 4407 | 0.15 | [0.14; 0.16] | 38.5% |
| **Random effects (BGLMM method)** | | | | | **10085** | **0.14** | **[0.13; 0.15]** | **−−** |
| **Random effects (HKSJ method)** | | | | | | **0.09** | **[0.00; 0.50]** | **100.0%** |
| Heterogeneity: $I^2 = 82\%$, $\tau^2 = 0.03$, $p < 0.01$ | | | | | | | | |
| **Actual code sharing rates** | | | | | | | | |
| Strcic 2022 | No restrictions | IPD | Manual | 16 | 888 | 0.02 | [0.01; 0.03] | 54.6% |
| Gkiouras 2020 | No restrictions | Paper | Manual | 1 | 35 | 0.03 | [0.01; 0.15] | 45.4% |
| **Random effects (BGLMM method)** | | | | | **923** | **0.02** | **[0.01; 0.03]** | **−−** |
| **Random effects (HKSJ method)** | | | | | | **0.02** | **[0.00; 0.05]** | **100.0%** |
| Heterogeneity: $I^2 = 0\%$, $\tau^2 = < 0.01$, $p = 0.68$ | | | | | | | | |

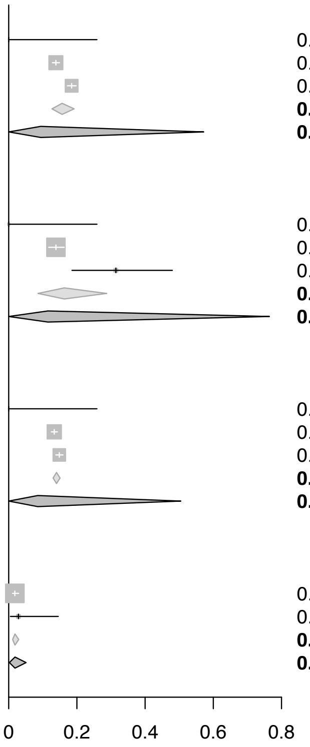

**Supplementary Table 1. Deviations from the original review protocol.**

| Original plan | Revised plan | Reason for modification |
| --- | --- | --- |
| Please refer to Table 1 in the review protocol (https://f1000research.com/articles/10-491/v2 - T1). | Two extra options were added to the 'risk of sampling bias' ("Sampled whole population of interest") and 'risk of article selection bias' categories ("No article screening performed"). Both items were considered low risk of bias. | The two options were added to account for meta-research articles that assessed all articles within a population of interest (e.g., assessed all articles indexed in PubMed Central a la Serghiou et al. (2021) [14]) or did not perform article screening respectively. |
| Prevalence rates will be transformed using the Freeman-Tukey double arcsine transformation and combined using standard inverse variance methods. | We pooled prevalence estimates by first stabilising the variances of the raw proportions using arcsine square root transformations, then applied random-effects models using the Hartung-Knapp-Sidik-Jonkman method. | Due to large sample size imbalances between included studies, and the negative influence such skewed ranges of sample sizes can have on the harmonic mean which is used to back-transform meta-analytic estimates transformed with the Freeman-Tukey double arcsine method [Schwarzer 2019], we decided to use the arcsine square root transformation instead. |
| We did not plan to conduct sensitivity analyses to investigate differences in pooled risk ratios when using generalized linear mixed models. | We examined differences in pooled risk ratios when using generalised linear mixed models to aggregate findings [32,33]. | We decided post-hoc to check the robustness of the meta-analyses of risk ratios when using bivariate generalised linear mixed effects models as proposed by Chu et al. (2012) [33]. Like the meta-analyses of proportions, we chose this method as it has been specifically recommended in situations when the event of interest is rare, and individual study sample sizes are small and circumvent the need to add arbitrary continuity corrections allowing the analysis of both single-zero and double-zero events [27; 165]. |
| We did not plan to collect information on data type, nor perform a subgroup analysis to explore its effects on the study's findings. | We collected data on data type and conducted a sub-group analysis to investigate whether prevalence estimates of public data sharing differed depending on the data type, or … | We decided to collect data on data type and perform a subgroup analysis exploring its effects on reported findings based on feedback from colleagues on the protocol. |

**Supplementary Table 1 continued. Deviations from the original review protocol.**

| Original plan | Revised plan | Reason for modification |
|---|---|---|
| We did not plan to conduct sensitivity analyses to investigate differences in pooled prevalence rates when excluding studies that used automated coding strategies. | We conducted sensitivity analyses to investigate differences in pooled prevalence rates when excluding studies that used automated coding strategies. | We decided to include this sensitivity analysis to evaluate whether pooled prevalence rates deviated when the results of meta-research studies which used automated coding strategies (methods have been shown to have inferior accuracy, positive predictive value (PPV), and negative predictive value (NPV) when compared to manual coding strategies) were removed. |
| We originally planned to perform subgroup analyses to explore differences in public data sharing frequencies between primary articles reporting the results of clinical trials or not, as well as articles reporting the results of studies using human participants versus not. | We conducted analyses of association for these outcomes rather than a subgroup analysis comparing pooled proportions between groups. Consequently, these two subgroup analyses have been included as secondary outcomes. | We changed the analysis plan for these outcomes due to the availability of data that allowed us to directly explore associations between both of these factors (i.e. calculation of risk ratios) |
| We originally planned to perform a subgroup analysis to explore differences in public data sharing frequencies between primary articles studying COVID-19 or not. | We conducted a sensitivity analysis to examine how data sharing frequencies changed when analyses were restricted to meta-research studies examining COVID-19. | We decided not to compare data and code sharing rates between COVID and non-COVID research because of the large amount of methodological heterogeneity in meta-research studies examining non-COVID research. |

**Supplementary Table 2. Risk of bias criteria.**

| Item | Low risk of bias | High risk of bias | Unclear risk of bias |
|---|---|---|---|
| Risk of sampling bias | The meta-research study evaluated a random sample of primary articles or sampled the entire population of interest. | The meta-research study included a non- or pseudorandom sample of primary articles. | The sampling frame for the sample of primary articles was unclear. |
| Risk of selective reporting bias | Eligible outcomes and associations reported in the protocol for the meta-research study were fully reported in the results section of the publication. | Not all eligible outcomes and associations reported in the protocol for the meta-research study were reported in the results section of the publication. | It was unclear if all eligible outcomes and associations were fully reported in the results section of the publication (e.g., because a study protocol for the meta-research study was unavailable). |
| Risk of article selection bias | Details about which studies were excluded from the study and why have been shared and match the criteria described in the methods, or no article screening needed to be performed (e.g., because all articles identified by a literature search were analysed) | Details about which studies were excluded and why were not reported. | Details about the eligibility criteria and study selection process was unclear. |
| Risk of errors in the accuracy of reported estimates | All outcome data were either manually coded by at least two people independently in parallel or coded by one person and checked in full by another. | Outcome data were manually coded by one researcher, an automated algorithm, or according to another methodology different from that outlined in the Low Risk category. | The method used to extract data from the included primary studies was unclear. |

**Supplementary Table 3. Findings of eligible meta-research studies where summary data were not available for the review (N=9).**

| Study | Year | Discipline | Journals examined | Primary study date range | Data types | Sample size | IPD available | Exclusion reason |
|---|---|---|---|---|---|---|---|---|
| Helliwell 2020b | 2020 | COVID-19,<br>MERS | Multiple | 2019-2020,<br>2018-2019 | Any | 398<br>55 | Partial | Reported prevalence estimates could not be coded in accordance with the study codebook |
| Hemkens 2016 | 2016 | General Medical | Multiple | 2012 | Clinical data | 124 | No | Reported prevalence estimates could not be coded in accordance with the study codebook |
| Jiao 2022 | 2022 | Multidisciplinary | PLOS One | 2014-2020 | Any | 127,935* | No | Prevalence estimates not reported separately for medical articles |
| McDonald 2017 | 2017 | General Medical | BMJ | 2015-2017 | Clinical data | 237 | Partial | Reported prevalence estimates could not be coded in accordance with the study codebook |
| Ramke 2018 | 2018 | Ophthalmology | Multiple | 2000-2014 | Clinical data | 153 | No | Prevalence estimates not reported |
| Rustici 2021 | 2021 | Biomedicine | Multiple | 2009-2013,<br>2012 | RNA-Seq,<br>Microarray | 1,114*,<br>347* | No | Prevalence estimates not reported separately for medical articles |
| Stodden 2018 | 2018 | Multidisciplinary | Science | 2009-2010 | Any | 204* | No | Reported prevalence estimates could not be coded in accordance with the study codebook and are also not reported separately for medical articles |
| Towse 202 | 2020 | Clinical Psychology | Multiple | 2014-2017 | Any | 1,900* | Partial | Prevalence estimates not reported separately for medical articles |
| Zhao 2017 | 2017 | Multidisciplinary | PLOS One | 2014-2015 | Any | 50* | No | Prevalence estimates not reported separately for medical articles |

**Supplementary Table 4. Meta-regression results.**

| | Model Coefficients | | | | | | Level 3 (Between-study) | | | Level 2 (Within-study) | | | | |
|---|---|---|---|---|---|---|---|---|---|---|---|---|---|---|
| | Intercept | SE | β* | SE | 95% CI | *p* | $\tau^2$ | $I^2$ | k | $\tau^2$ | $I^2$ | o | AIC | BIC |
| **Declared data sharing** | | | | | | | | | | | | | | |
| - Three-level (All) | -18.4290 | 1.4417 | 0.0093 | 0.0007 | 0.0078-0.0107 | <0.0001 | 0.0124 | 90.66% | 27 | 0.0013 | 9.11% | 155 | -278.92 | -266.80 |
| - Three-level (Manual) | -34.0273 | 10.1446 | 0.0170 | 0.0050 | 0.0070-0.0270 | 0.0010 | 0.0117 | 55.06% | 25 | 0.0032 | 15.05% | 118 | -149.50 | -138.48 |
| - Two-level (All) | -17.9399 | 2.4845 | 0.0090 | 0.0012 | 0.0066-0.0115 | <0.0001 | 0.0084 | 99.62% | 155 | - | - | - | -195.29 | -186.20 |
| - Two-level (Manual) | -42.7527 | 8.9317 | 0.0213 | 0.0044 | 0.0126-0.0301 | <0.0001 | 0.0132 | 67.64% | 118 | - | - | - | -114.37 | -106.10 |
| **Actual data sharing** | | | | | | | | | | | | | | |
| - Three-level (All) | -9.0520 | 6.5014 | 0.0045 | 0.0032 | -0.0018-0.0109 | 0.1615 | 0.0088 | 76.38% | 26 | 0.0002 | 2.06% | 125 | -230.02 | -218.77 |
| - Three-level (Manual) | -16.0684 | 8.5307 | 0.0080 | 0.0042 | -0.0004-0.0164 | 0.0604 | 0.0087 | 61.08% | 25 | 0.0004 | 2.54% | 119 | -205.38 | -194.33 |
| - Two-level (All) | -20.4047 | 6.9122 | 0.0102 | 0.0034 | 0.0034-0.0170 | 0.0036 | 0.0057 | 69.65% | 125 | - | - | - | -197.37 | -188.93 |
| - Two-level (Manual) | -21.0863 | 7.5362 | 0.0105 | 0.0037 | 0.0031-0.0179 | 0.0058 | 0.0065 | 55.57% | 119 | - | - | - | -178.70 | -170.42 |
| **Declared code sharing** | | | | | | | | | | | | | | |
| - Three-level (All) | -7.1880 | 0.6262 | 0.0036 | 0.0003 | 0.0030-0.0042 | <0.0001 | 0.0010 | 82.70% | 24 | 0.0002 | 15.03% | 139 | -428.82 | -417.14 |
| - Three-level (Manual) | -1.5145 | 10.6961 | 0.0008 | 0.0053 | -0.0098-0.0113 | 0.8852 | 0.0014 | 18.54% | 22 | 0.0000 | 0% | 102 | -236.20 | -225.78 |
| - Two-level (All) | -4.4004 | 0.7886 | 0.0022 | 0.0004 | 0.0014-0.0030 | <0.0001 | 0.0005 | 94.92% | 139 | - | - | - | -400.39 | -391.63 |
| - Two-level (Manual) | -5.4033 | 11.2150 | 0.0027 | 0.0056 | -0.0083-0.0137 | 0.6290 | 0.0009 | 13.19% | 102 | - | - | - | -235.61 | -227.80 |
| **Actual code sharing** | | | | | | | | | | | | | | |
| - Three-level (All) | -5.1997 | 10.5109 | 0.0026 | 0.0052 | -0.0078-0.0129 | 0.6199 | 0.0009 | 15.15% | 21 | 0.0000 | 0% | 99 | -242.96 | -232.66 |
| - Three-level (Manual) | NA | NA | NA | NA | NA | NA | NA | NA | NA | NA | NA | NA | NA | NA |
| - Two-level (All) | -11.2948 | 10.6347 | 0.0056 | 0.0053 | -0.0048-0.0161 | 0.2894 | 0.0006 | 10.26% | 99 | - | - | - | -241.18 | -233.45 |
| - Two-level (Manual) | NA | NA | NA | NA | NA | NA | NA | NA | NA | - | - | - | NA | NA |

**Supplementary Table 5. Sensitivity analyses for secondary outcomes and subgroup analyses.**

| | Declared public sharing | | | | | Actual public sharing | | | | |
|---|---|---|---|---|---|---|---|---|---|---|
| | RR | 95% CI | 95% PI | k | $I^2$ | RR | 95% CI | 95% PI | k | $I^2$ |
| **Association between data and code sharing** | | | | | | | | | | |
| - HKSJ method | 8.03 | 2.86-22.53 | 0.33-194.43 | 12 | 32% | 42.05 | 12.15-145.52 | 0.94-1879.62 | 7 | 0% |
| - BGLMM method (SZC) | 7.88 | 2.44-18.01 | NA | 12 | NA | 40.42 | 15.45-120.39 | NA | 7 | NA |
| - BGLMM method (DZC) | 10.51 | 3.00-18.01 | NA | 23 | NA | 52.85 | 9.46-132.52 | NA | 17 | NA |
| - Low ROB | - | - | - | - | - | - | - | - | - | - |
| - FAIR studies | 11.84 | $0-1.33\times10^{7}$ | NA | 2 | 82% | - | - | - | - | - |
| - IPD only | - | - | - | - | - | - | - | - | - | - |
| - Manual coding | 4.52 | 1.38-14.86 | 0.20-101.05 | 10 | 0% | - | - | - | - | - |
| **Trial versus non-trial** | | | | | | | | | | |
| - HKSJ method | 0.69 | 0.45-1.07 | 0.12-4.13 | 23 | 0% | 0.96 | 0.53-1.72 | 0.15-5.95 | 19 | 0% |
| - BGLMM method (SZC) | 0.55 | 0.35-0.77 | NA | 23 | NA | 0.67 | 0.26-1.39 | NA | 19 | NA |
| - BGLMM method (DZC) | 0.56 | 0.37-0.79 | NA | 25 | NA | 0.69 | 0.27-1.52 | NA | 24 | NA |
| **Human versus non-human** | | | | | | | | | | |
| - HKSJ method | 0.65 | 0.42-0.99 | 0.12-3.61 | 19 | 57% | 0.44 | 0.24-0.81 | 0.05-3.57 | 16 | 28% |
| - BGLMM method (SZC) | 0.69 | 0.46-1.01 | NA | 19 | NA | 0.58 | 0.29-1.00 | NA | 16 | NA |
| - BGLMM method (DZC) | 0.69 | 0.48-1.00 | NA | 20 | NA | 0.59 | 0.30-0.97 | NA | 20 | NA |

SZC – Did not include studies with no events in both groups in analyses, DZC – Included studies with no events in both groups in analyses
